# Advances and limitations of artificial intelligence-assisted identification of pathogenic fungi

**DOI:** 10.1101/2025.07.16.25331671

**Authors:** J. Benjamin Stielow, Sarah Ahmed, G. Sybren de Hoog

## Abstract

**Objectives:** We developed and tested multiple computer-vision image classifiers, for their ability to identify a large set of common and rare pathogenic molds. Aim of the study was to create a comprehensive global benchmark towards the novel, emerging field of computer vision driven diagnostics of pathogenic microbes. If successfully implemented, a high-resource clinical setting could greatly benefit from this adjunct technique to supplement molecular sequencing and mass spectrometry driven methodologies, while in a low resource setting, it could provide enormous possibilities to enhance diagnostic precision in rural and remote geographies.

**Methods:** We selected 114 representative fungal pathogens represented by 123 strains obtained from the unique images implemented within the ‘Atlas of Clinical Fungi’, to serve as core dataset. The image classifiers were designed with a rigorous testing and evaluation strategy, at a yet unprecedented level of detail. We designed the framework, within the TENSORFLOW environment, testing multiple transfer-learning approaches, as well hybrid architectures comprising both, features of convolutional neural networks (CNN) and advanced vision transformers (ViT).

**Results:** We achieved a global identification accuracy of > 88% for the validation partition with our best model (Test accu. 87%, Train. accu. 96%). Simulations indicated that extended training time would lead to further accuracy improvements, particularly with greater data richness. Our results also highlight complex de-black-boxing approaches in interpreting image classification, never shown for microbial computer vision diagnostics to date.

**Discussion:** Besides quantitative limitations of representative strains per tested species, our approach reflects a significant scientific novelty to the field. While tested mainly on molds and a small subset of common bacteria as a control set, the methodology is universally applicable to yeasts and bacteria rendering the technique attractive for future diagnostics in the clinical setting.

## Introduction

In clinical settings, accurate and prompt identification of fungal pathogens is vital for disease diagnosis, guiding antifungal therapy, and effective treatment (Cornely *et al*. 2012; Giannella *et al*. 2025). Current diagnostic methods—relying on molecular techniques and mass spectrometry, such as MALDI-ToF and LC-MS—are restricted to resource-rich clinical settings and prone to human error, highlighting a need for advanced, automated low-cost solutions (Torres-Sangiao *et al*. 2021; Wu & Gadsden 2023) for independent verification.

Deep learning, particularly Convolutional Neural Networks (CNNs), has revolutionized medical imaging and pathology. By automatically learning hierarchical features from raw data, CNNs excel at classifying both macro- and micro-morphological images of microbial pathogens (Pawłowski *et al*. 2022; Tsang *et al*. 2025). Such AI-driven diagnostic approaches can match or exceed the accuracy, consistency, and speed of conventional methods—while dramatically reducing costs. Recent CNN implementations employing transfer learning from large datasets like ImageNet have demonstrated remarkable performance even with limited domain-specific data. Moreover, integrating attention mechanisms refines these models by focusing analyses on diagnostically relevant image regions and suppressing background noise (Ma *et al*. 2021; Huang *et al*. 2023; Gumus 2024; Rawat *et al*. 2024).

Despite these advances, a significant challenge remains in the scarcity of annotated, high-quality image data. Annotating fungal images is labor-intensive and requires expert knowledge, making large-scale data collection time-consuming. Modern CNN architectures—such as ConvNeXtTiny— (Liu et al. 2022) have been employed alongside transfer learning frameworks that incorporate transformer-inspired components. These transformer architectures, with their self-attention mechanisms (Thirunavukarasu & Kotei 2024; Hörst et al. 2024; Halder et al. 2024).have a significant impact on model performance by capturing long-range dependencies and complex patterns, thereby enhancing feature extraction. By leveraging these advanced design principles and augmenting training data (Shorten & Khoshgoftaar 2019; Yang et al. 2023) through synthetic generation, deep learning models can achieve high classification accuracy demonstrating strong potential to complement traditional diagnostics that often have limited accuracy in filamentous fungal identification.

Rani et al. (2025) published a recent computer vision application in microbial imaging, and more specifically to general mycology. The authors developed a time-lapse imaging framework combining ResNet50 backbones with Vision Transformer modules, classifying over 110 fungal strains with > 90 % accuracy within 24 hours of incubation—reducing diagnostic turnaround by multiple days. The FungiCLEF25 challenge (https://www.kaggle.com/competitions/fungi-clef-2025/) show cased few-shot learning: prototypical networks achieved > 85 % accuracy across 200 mushroom taxa with as few as 3–5 images per species, demonstrating feasibility in resource-limited environments.

On the horizon and one of the most recent top-end publications is Microsoft’s Medical Superintelligence concept (Microsoft 2025; https://microsoft.ai/new/the-path-to-medical-superintelligence/) envisions a multi-agent system of specialized (GPT) AI “assistants,” where one agent performs image analysis, another processes clinical metadata, and a third synthesizes treatment recommendations. The concept is collectively achieving diagnostic accuracy up to four-fold higher than human clinicians, as demonstrated in over a broad set of NEJM case studies. Adapted to mycology, such orchestration could automate the full spectrum of fungal diagnostics, from plate image interpretation, over genomic data to antifungal susceptibility inference.

However, translating these innovations into clinical practice requires adherence to trustworthy AI principles. The FUTURE-AI framework (Weber et al. 2023) prescribes transparency, fairness, robustness, and ethical alignment—imperatives for patient safety and equitable access to AI-driven fungal diagnostics.

Building on these cutting-edge developments, our more than two-year study integrates state-of-the-art CNN-transformer hybrids with temporal imaging, into a unified pipeline for filamentous fungal classification using standard macro-morphological images. Such comprehensive AI frameworks utilizing computer vision approaches may deliver on rapid, transparent, and clinically actionable diagnostics. Rapid recognition of fungi by colony appearance is not only helpful under conditions of limited resources, and for independent verification of the identity of diagnosed strains.

## Material and methods

### Ethics

Ethical permission for this research was not required as it was undertaken on stored clinical isolates, without reference to patient identifiable data.

### Fungal identification, culturing and image acquisition

High-resolution images of cultures originated from the Atlas of Clinical Fungi encyclopedia project (www.atlasclinicalfungi.org) and were collected over a period of the past 10+ years. All strains were type or reference strains and are hosted at the Westerdijk Fungal Biodiversity Institute (former CBS-KNAW), Utrecht, the Netherlands. Strains were grown on a wide variety culture media and photographed with a Canon EOS 600D in standardized bright-white lightening (single illumination condition), at a single top-front facing 90° angle, and images were stored in sRGB, JPG format at 5-mega pixel resolution. Most strains were grown on at least two culture media, to depict morphological variability and to mimic changing culturing condition (Table 1). The total evaluated dataset contained 853-curated raw JPG images, with front and backside captures, consisting of 114 fungal species represented by 123 strains. Of those 123 strains, nine species were grown at two separate time points (Total sum 132 classes and conditions). Molecular and mass spectrometry associated methods (DNA barcoding/MALD-ToF) were applied to pre-identify strains and defining the reference method in order to associate species labels to strains (Performed by Westerdijk FBI). Examples of cultured species and raw input image data is provided in figure 1.

**Figure 1.**
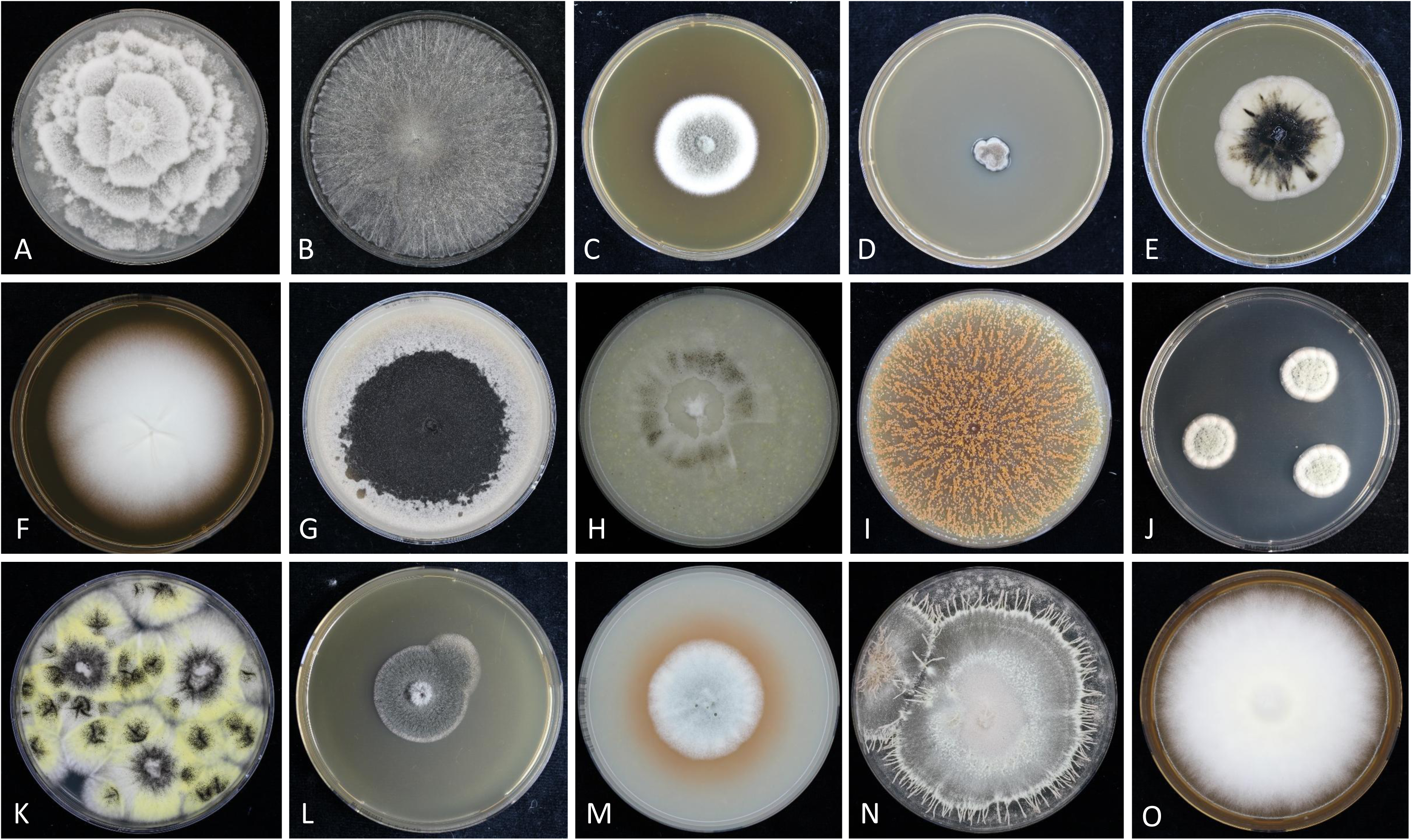
Representative images of studied fungal pathogens included in the dataset. Species shown are: A, *Mortierella wolfii*; B, *Papulaspora equi*; C, *Paraconiothyrium cyclothyrioides*; D, *Exophiala salmonis*; E, *Aureobasidium pullulans*; F, *Lophophyton gallinae*; G, *Mycocentrospora acerina*; H, *Sporothrix stenoceras*; I, *Thermoascus crustaceus*; J, *Penicillium capsulatum*; K, *Aspergillus awamori*; L, *Bipolaris australiensis*; M, *Myxotrichum deflexum*; N, *Paecilomyces fumosoroseus*; O, *Microsporum canis*.

**Table 1.**
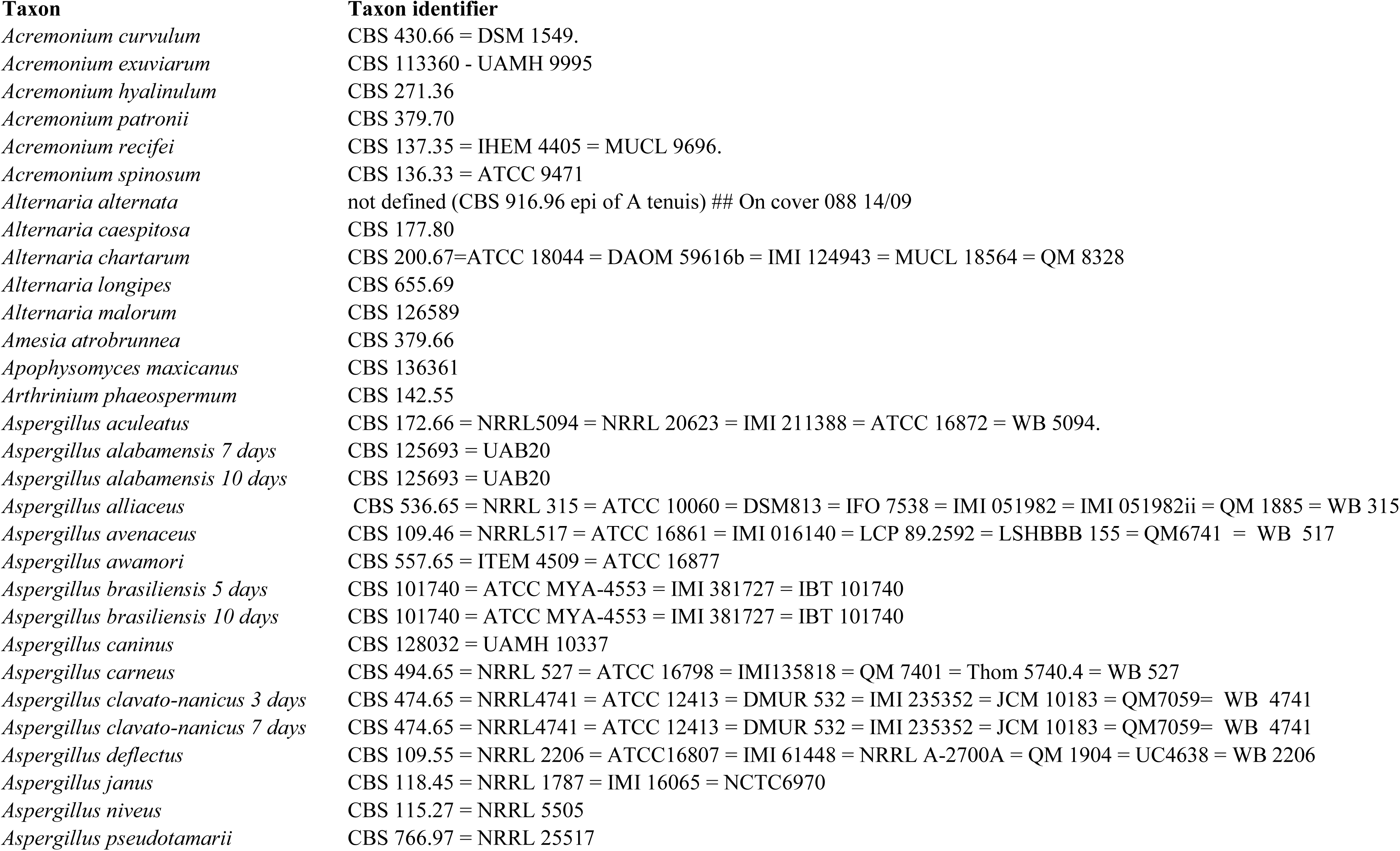
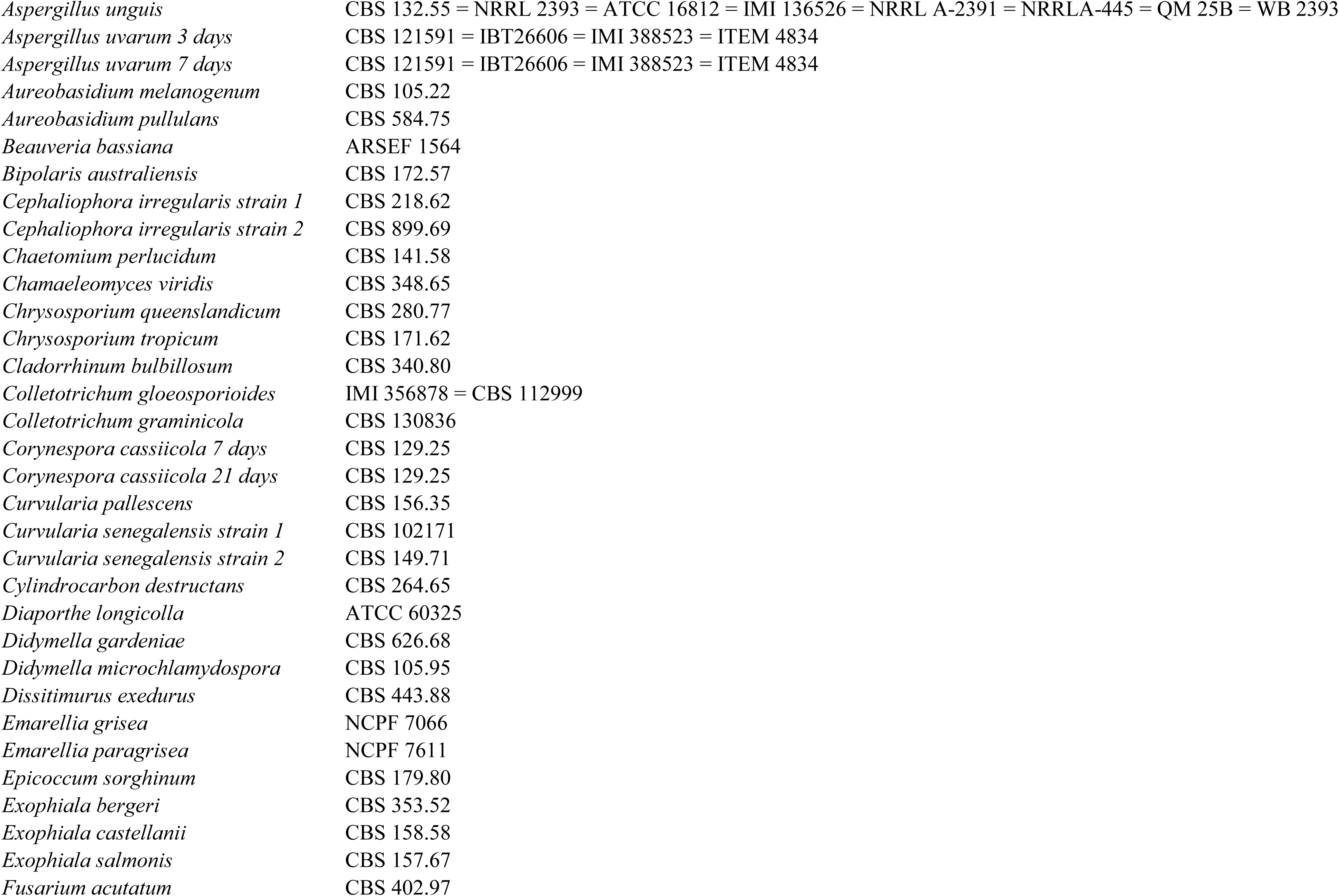
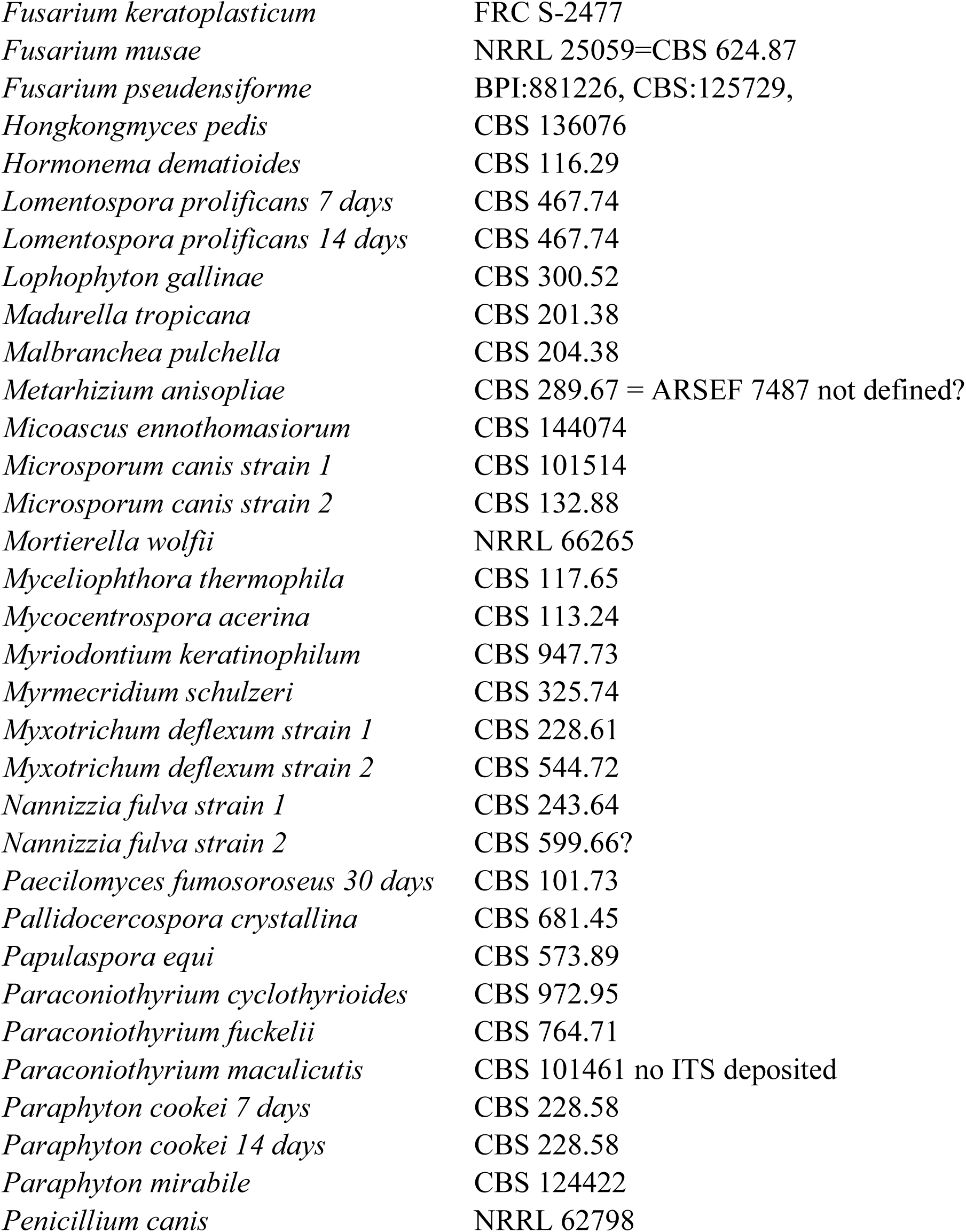
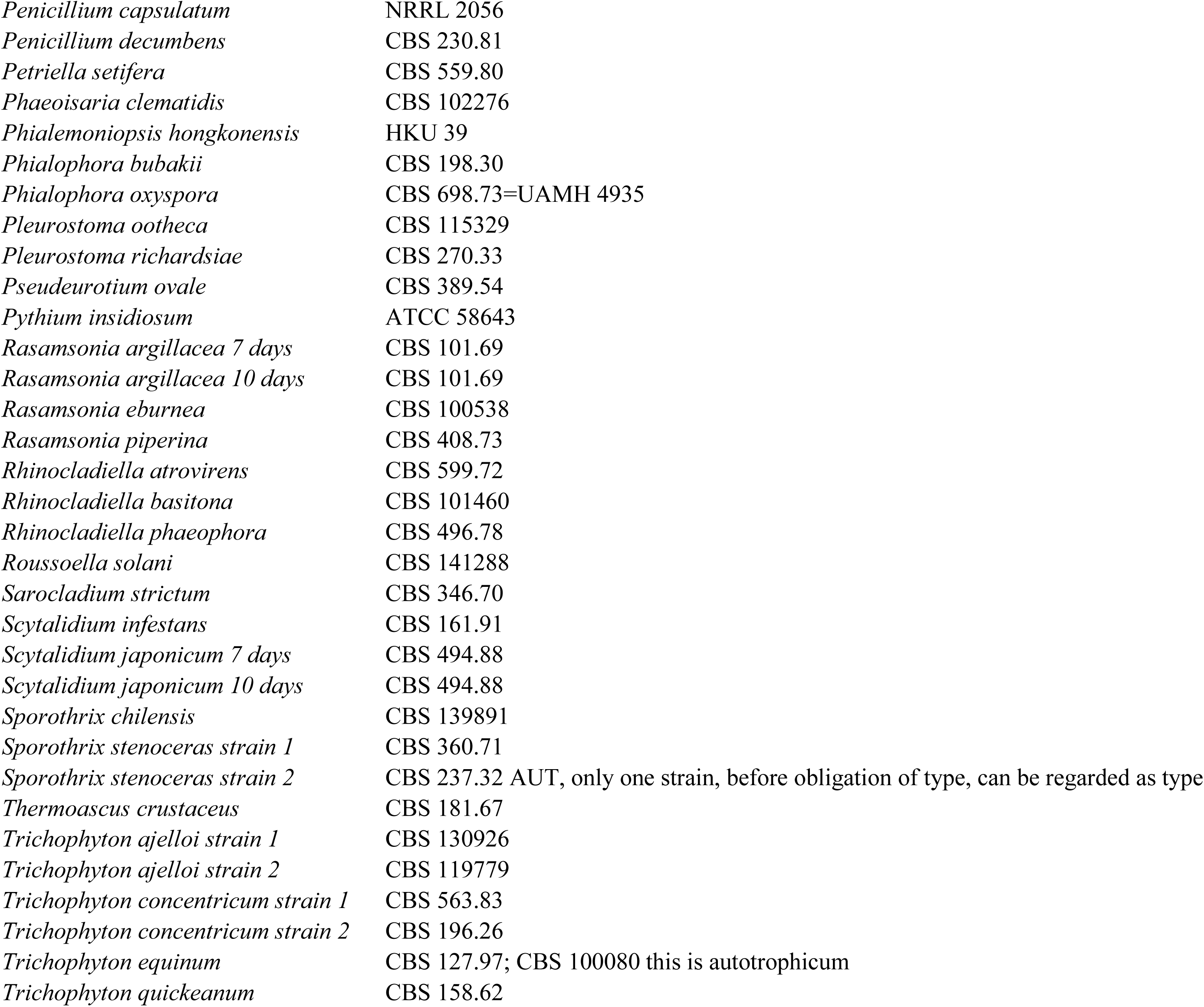
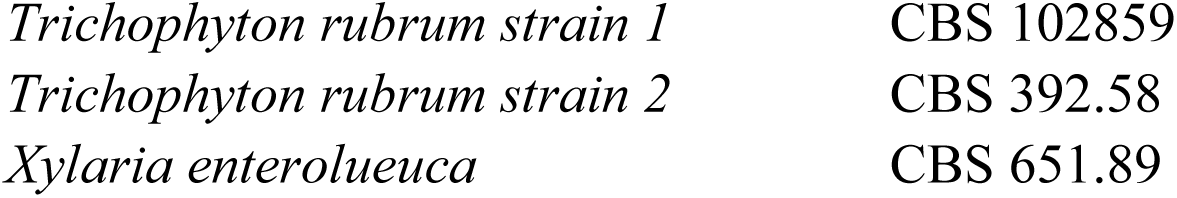
Reference strains and corresponding metadata utilized in the current study.

### Compute resources, initial study design and benchmark

Our compute resources comprised various high performance instances to conduct the described diagnostic simulations, i.e., a Dell PowerEdge R750XA, with 38 cores, 4 TB RAM and 2 Nvidia Ampere A100 80 GB HBM2E units or alternatively, instances with similar specifications were used as webhosted services on Google Cloud Platform (GCP).

### Methodological description initial study design and benchmark

Initially, we developed a CNN base model within the popular TENSORFLOW environment, and using associated automation procedures to record statistical testing metrics. Importantly, we conducted an ablation study that scaled up the model’s complexity to determine its optimal setup given the image input data as well applied, to increase image diversity, a standardized augmentation technique.

Briefly, the ablation study and the benchmark against the Neurosys AGAR dataset outlines a complete end-to-end deep learning computer vision pipeline for image classification with several key optimization phases that were relevant to the later stages of the study, these are: data processing, augmentation, model construction, training, and evaluation. In the first stage, the code imports essential libraries for linear algebra, data visualization, and machine learning—including NumPy, Pandas, Seaborn, Matplotlib, and Scikit-learn—and additional modules for balancing imbalanced datasets (from imblearn) and building deep learning models (from TensorFlow Keras).

The data preparation section begins with defining helper functions to generate a dictionary mapping each class name (derived from subdirectory names in the input data folder) to a numeric label. It then counts the available training images for each class using file globbing, and flags classes with fewer than a defined threshold of images. Once the class structure and image counts are established, the code proceeds with a function to load and preprocess images: each image is read, resized to a standard dimension (256×256), converted to an array, and the corresponding label is one-hot encoded. This step ensures a consistent dataset structure for later training.

Additionally, the script also integrates image data augmentation using TensorFlow’s ImageDataGenerator. This augmentation process involves applying random transformations such as rotations, flips, and zooms to increase the diversity of the training dataset, particularly for under-represented classes to the original raw images. The augmented images are saved locally and later reloaded, resized, and re-split into training, validation, and test sets. These steps collectively maximize data variability and contribute to model robustness.

As described in the main text body, the raw dataset consisted of 853-curated images. Depending on the experimental simulation condition (Table 2), we diversified these initial images by a simple augmentation principle introducing rotations, flips and random zooming. We ad-hoc specified three test sizes with 20, 50 and 80 augmented images per class, which subsequently, resulted in 2640 (dataset_size 20), 6600 (dataset_size 50) and 10560 (dataset_size 80) additional images for the initial classification study. These quantities remained identical between different simulations, given model complexity up scaling (ablation).

**Table 2.**
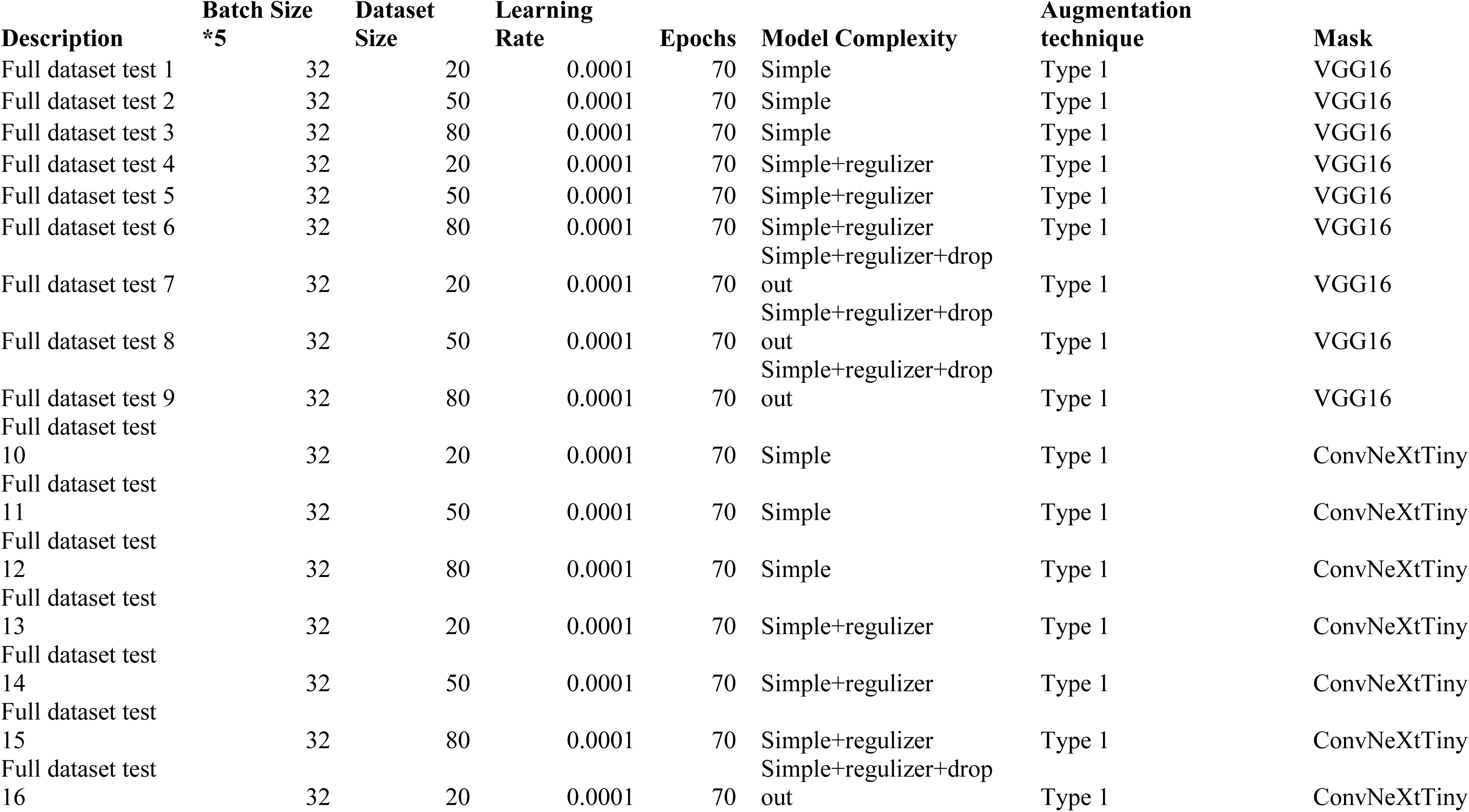
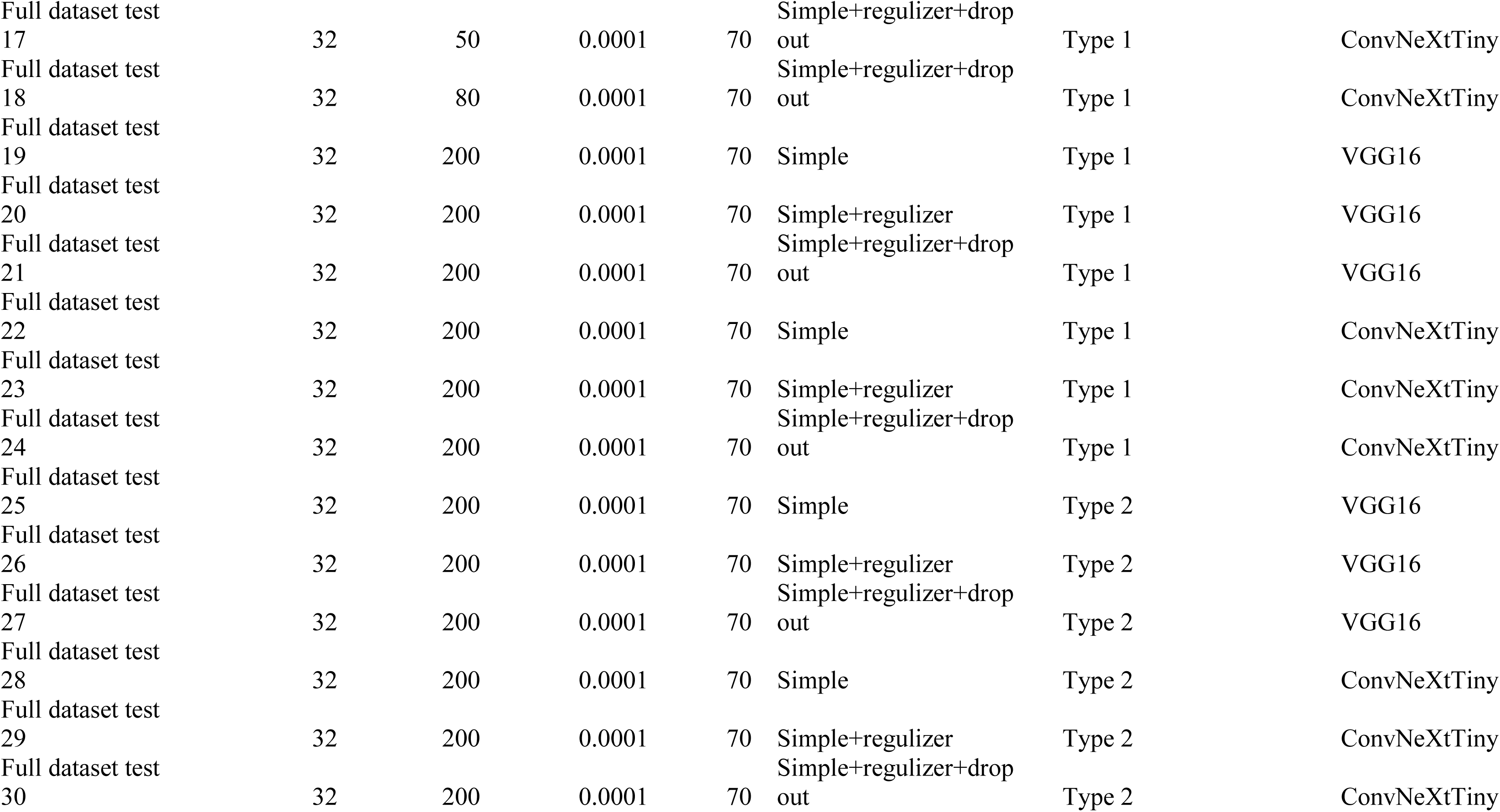
Technical summary of initial pipeline development and ablation study parameters.

The second segment of the code transitions into building and evaluating a Convolutional Neural Network (CNN) architecture using the pre-trained VGG16 and ConvNeXtTiny model as the base. The model construction leverages transfer learning by freezing the base layers (through a separate function to set layers as non-trainable) and appending additional layers—including flattening, dense layers with ReLU activations and L2 regularization, dropout layers for overfitting prevention, and a final softmax prediction layer—to tailor the architecture for the specific image classification task.

We iterated the simulation over a fixed batch-size range of 32*2 in five iterations (VGG16) and three iterations (ConvNeXtTiny) each, at a fixed learning rate, a fixed number of epochs, varying quantity of images as input (Table 2), a single augmentation type and two transfer learning backbones (VGG16 and ConvNeXtTiny). In total we conducted seventy-two simulations within the ablation study to determine the initial CNN base model performance.

Custom performance metrics such as precision, recall, and an F1 score are defined to monitor and evaluate the model’s learning during training. The model is then compiled using the Adam optimizer with a predefined learning rate, and a categorical crossentropy loss function is set due to the multi-class nature of the classification task.

The training process is further enhanced by including callbacks like EarlyStopping (to restore the best weights and prevent overfitting) and ModelCheckpoint (to save the model that achieves the highest validation accuracy). The training procedure accounts for class imbalance by computing sample weights based on the balanced class distribution, ensuring that under-represented classes have a proportional influence on the loss function. Finally, the code generates visual outputs: it plots the training history—visualizing loss, accuracy, precision, recall, and F1 scores over epochs—and creates confusion matrices to assess classification performance on the test set. In addition, the code integrates with OpenPyXL to create and update an Excel workbook that logs key training parameters, evaluation metrics, and visualizations such as history plots and confusion matrices across multiple training iterations with varying batch sizes. This combination of data preparation, augmentation, model development, and comprehensive evaluation constitutes a robust pipeline aimed at improving classification performance, ensuring reproducibility, and documenting experiments effectively.

In addition, we conducted; a benchmark study utilizing the independent Neurosys AGAR dataset (https://agar.neurosys.com/), which we simulated with the same technical methodology as described above. This initial source dataset contained 200 images each, for the microbial species *Bacillus subtilis, Escherichia coli, Pseudomonas aeruginosa, Staphylococcus aureus* and *Candida albicans*. We applied the simple and more sophisticated augmentation methodology to diversify the dataset. The concept paper is available at https://arxiv.org/abs/2108.01234.

An example for the augmented data is obvious from figure 2. A-J indicates various growth stages of *Aspergillus awamori*, as well front and backside captures for exemplified original images that underwent augmentation. Original images, as of figure 1, contained a black static background.

**Figure 2.**
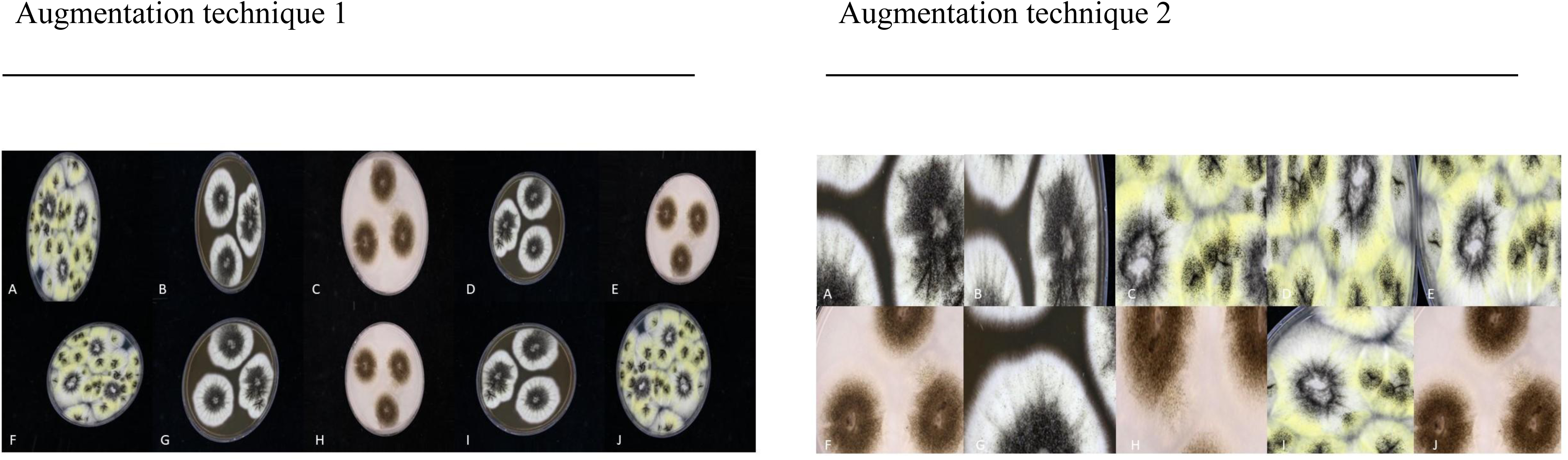
Examples of simple and advanced image augmentation strategies for synthetic data generation. Panels A–J depict various growth stages of *Aspergillus awamori*, including both front and back colony images used for augmentation. Augmented images employed in classification (excluding those in the ablation study) feature backgrounds removed and emphasize authentic colony morphology.

### Methodological description of final classifier design

The resulting performance metrics from the base model design served as findings to design the final model and its classification head. This included the design of a second, more sophisticated custom image generator to increase precision of augmented images as well their resolution, since the ablation study was designed on a simpler augmentation principle to solely overcome the quantitative and balancing limitations of the original dataset and subsequently the initial model design (i.e., low number of images per class). This second augmentation principle was mainly implemented to maximize information density and to reduce noise, such as biases by lowering surface of structures that are non-fungal (such as parts of a petri dish and agar).

Data were loaded and preprocessed using a computer vision pipeline implemented in Python that combined standard libraries (eg. NumPy, Pandas, Matplotlib) with highly advanced deep learning and augmentation frameworks (TensorFlow Keras, imbalanced-learn, and custom augmenters). First, the class names were dynamically extracted from a specified directory hierarchy via a custom function, ensuring that class labels were indexed starting at zero. Individual images were then loaded using the Python Imaging Library (PIL), resized to a predetermined square dimension (1024 × 1024 pixels), and converted into arrays. A one-hot encoding transformation was applied to the class labels using TensorFlow, ensuring that the number of output classes was correctly set.

Very specific attention was devoted to preventing data leakage throughout the pipeline. Initially, the raw image dataset was partitioned into training, validation, and testing subsets using stratified sampling to preserve the class distribution. The splitting process was performed in two steps using Scikit-Learn’s train_test_split with a fixed random seed of 42 for reproducibility. First, the dataset was divided into 60% training data and 40% temporary data. The temporary partition was then equally subdivided into validation and test sets (i.e., 20% each of the original data). This multi-step approach ensured that there was no overlap between training, validation, and testing data.

To further improve model robustness and address class imbalance, data augmentation was applied. A custom image data generator—extending Keras’s ImageDataGenerator—was implemented to perform randomized precision zoom transformations up to 50% (while preserving the exact aspect image ratio of the source image), horizontal and vertical flips, up to 10% width and height shifts, and appropriate gap filling. This augmentation process was specifically designed to synthetically expand the number of images in classes with fewer samples to meet a predefined minimum threshold (e.g., 40 images per class in the final simulations). Once augmented, these synthetic images were stored separately, and subsequently loaded into an augmentation dataset.

The overall training pipeline incorporated both the original and augmented data. First, the original dataset was split into training, validation, and test partitions as described. Independently, the augmented data underwent a similar stratified splitting process. Finally, the training sets from the original and augmented datasets were concatenated to create a combined training set, and similarly for the validation and test sets. After concatenation, each combined dataset was shuffled using a fixed random seed to ensure consistency and reproducibility. This combined and shuffled dataset was then normalized via a custom preprocessing function before being fed to the convolutional neural network. If augmentation caused imbalances, a further stratified split rebalanced the classes, and each partition was independently shuffled to prevent data leakage.

This careful and layered data handling strategy, which preserves class distributions and avoids overlap between training, validation, and test sets, was crucial to minimizing data leakage and optimizing the model’s generalizability.

The study was based on different fine-tunable architectures (VGG16, ConvNeXtTiny, Efficientnet V2b2 and Densenet 121) and pre-trained on ImageNet. In our implementation, the majority of the base model’s layers were frozen to leverage the pretrained weights; however, several convolutional layers were explicitly marked for potential visualization (e.g., via Grad-CAM) and later unfrozen during fine-tuning. A custom hypermodel class was defined to support hyperparameter optimization using Keras Tuner’s RandomSearch algorithm. Within this class, several dense layers were appended to a global average pooling layer, with hyperparameters (such as the number of units, dropout rates, and learning rates) dynamically sampled within predefined ranges. This model was then compiled with the Adam optimizer and custom metrics (accuracy, precision, recall, and a custom F1 score) implemented with TensorFlow backend functions to accurately reflect the performance on imbalanced datasets.

For model evaluation and validation, k-fold cross-validation (with k = 5) was employed. The KFold function from Scikit-Learn was used with shuffling and a constant random seed to generate reproducible folds. Within each fold, the training data was further partitioned into training and validation subsets, ensuring no overlap between these sets to avoid data leakage. The hyperparameter tuning step was performed on the training splits, and a separate early stopping criterion along with model checkpoints was applied during fine-tuning to prevent overfitting. Metrics were recorded from both the tuning and final training phases, and the performance of the model on the reserved test set was evaluated using confusion matrices, and additional custom metrics to supervise model performance in real time.

Finally, all experimental results were automatically logged and exported to individual Excel workbooks and csv files for subsequent analysis. Plotting functions were used to visualize model history (loss, accuracy, F1 score, precision, and recall over epochs) and confusion matrices. A comprehensive evaluation is available in data s1.

The benchmark study conducted utilizing the Neurosys AGAR dataset as described above, was not repeated a second time with the final classifier design.

Precision augmentation, as of the second methodology outlined in the section final classifier design, is showcased in figure 2.

### Visual classification performance and image evaluation

One of the technical hallmarks of this study design, was the implementation of an end-to-end solution, specifically targeting insights into the inner decision-making processes of a deep convolutional network we projected high-dimensional feature representations from different network layers into a 2D space using supervised UMAP (Uniform Manifold Approximation and Projection). Complementing this, Grad-CAM (Gradient-weighted Class Activation Mapping) and guided back propagated Grad-CAM techniques were employed to generate intuitive image heat maps that indicate which regions of the input images influence the model’s predictions most significantly. These visualization techniques help in diagnosing model behavior, ensuring its reliability and transparency.

Initially, the pipeline standardizes image preprocessing by loading and resizing images to a uniform size of 1024 × 1024 pixels and normalizing pixel values to a [0, 1] range, thus ensuring consistent input quality for further processing. Once preprocessed, images are passed to the model for feature extraction; a dedicated function extracts neural activations from user-specified layers via a truncated model, enabling the isolation of intermediate representations that capture varying levels of abstraction—from low-level complex geometric edge detections among fungal colonies in initial layers to high-level semantic features in deeper layers. These high-dimensional features are subsequently reduced to a two-dimensional space using Uniform Manifold Approximation and Projection (UMAP), which preserves both local neighborhood structures and global relationships, thereby facilitating the visualization of feature clusters and class separability. The resulting UMAP projections, are rendered using a consistent colormap and saved as PDFs. UMAPs provide a clear macro-level overview of how the network differentiates among classes given the respective abstraction level of the analyzed layer, thus highlights areas of potential overlap or ambiguity in the learned embeddings.

In parallel, the pipeline employs ‘Gradient-weighted Class Activation Mapping’ (Grad-CAM) along with guided Grad-CAM to generate heatmaps that visually explain the model’s predictions; by computing gradients with respect to activations in chosen convolutional layers, Grad-CAM produces a coarse localization map that underscores the regions of an input image, which are most influential for a particular prediction. While guided Grad-CAM refines these maps naturally, by merging them with guided backpropagation, the obtained maps are resulting in sharp and more detailed visualization of the features driving the predictive decision process.

These interpretability high-end computer vision techniques are essential for diagnosing model behavior, verifying that the network focuses on relevant image regions, and ultimately building trust in the predictive system by revealing both the global structure of its latent space and the local, pixel-wise contributions to its decision-making. The entire pipeline is structured to facilitate both macro-level analyses of feature distributions via UMAP and micro-level analyses of model predictions through Grad-CAM based methods. These techniques are providing a powerful tool to understand, debug, and validate deep learning models in a highly rigorous and transparent manner.

### Quantitative assessment of uncertainty

To systematically assess classifier confidence and output uncertainty, we extracted three quantitative metrics from the softmax output vector of each model for every test-set prediction: (1) Prediction Probability, defined as the maximum softmax value across classes, representing the model’s confidence in its most probable label; (2) Entropy, calculated as the Shannon entropy of the predicted class distribution, providing a scalar measure of overall prediction uncertainty for each sample; and (3) Top-2 Margin, computed as the absolute difference between the highest and second-highest softmax probabilities, which reflects the model’s decisiveness between the two most likely classes. These metrics were implemented in Python using NumPy and pandas, and computed as part of the evaluation pipeline following inference on the test data. Each metric was exported to a dedicated CSV files for further analysis, with distributions visualized as histograms using Matplotlib. This approach enabled a comparative assessment of prediction calibration, model sharpness, and class-separation confidence across different CNN architectures and experimental conditions. The entropy and top-2 margin provided complementary perspectives: while entropy captured both confident and diffuse predictions, the top-2 margin specifically highlighted cases of ambiguous class competition. By analyzing the joint and marginal distributions of these metrics, we were able to distinguish between confidently correct, confidently incorrect, and uncertain predictions, informing model calibration and reliability analyses across all evaluated backbones.

## Results

To rigorously assess capacity for fungal-colony classification under data scarcity, we conducted an ablation study over three augmented training-set sizes—20, 50, and 80 images per species— while holding all other methodological factors constant (Data S1). Augmented images were generated via random rotations, flips, and zooms; preprocessed to 256 × 256 pixels; and split into stratified training, validation, and test partitions as described previously. The classification head comprised a global-average-pooling layer followed by two dense layers (each with ReLU activation), a 30 % dropout, and a final softmax layer; all experiments used categorical cross-entropy loss, the Adam optimizer with a fixed learning rate of 1 × 10⁻³, and EarlyStopping (patience = 10 epochs on validation loss). To probe the effect of complexity control, we compared three variants: the base head (“simple”), the head with L₂ weight decay (λ = 1 × 10⁻⁴) on dense weights (“+reg”), and the head with both L₂ regularization and 30 % dropout (“+reg+dropout”). Violin plots of accuracy, F1-score, recall, precision, and loss summarized performance across five cross-validation folds for each model variant.

### Initial study design and benchmark (NeuroSYS control dataset)

To test model robustness and domain transferability, we benchmarked both our classical (VGG16) and modern (ConvNeXtTiny) convolutional backbones against the Neurosys AGAR dataset, which serves as an external control due to its predominantly bacterial rather than fungal image content. This deliberate domain shift, involving different colony morphologies, textures, and image backgrounds, provides a stress test for deployment.

On the Neurosys AGAR control dataset, the VGG16 backbone, when equipped with a simple head, achieved median test and validation accuracies of approximately 89% and 90%, with F1-scores consistently above 0.90. L₂ regularization modestly improved both performance and interfold stability, raising median accuracies to 92% and reducing variance across splits. However, introducing both L₂ regularization and dropout led to marked instability and significant declines in recall and F1, with distributions becoming highly bimodal or even trimodal. This suggests that, when confronted with a domain mis-match, classical CNNs are prone to underfitting and suffer from poor calibration, likely because their learned filters overemphasize dataset-specific backgrounds and global contrasts rather than truly diagnostic morphological features.

By contrast, ConvNeXtTiny demonstrated stronger and more stable performance under the same conditions. Across five cross-validation folds, ConvNeXtTiny’s simple classification head achieved median test and validation accuracies of ∼95% and ∼96%, with macro-F1 scores around 0.94. The application of L₂ regularization and dropout did not degrade performance, but rather helped maintain robust classification across the varied control samples, with narrow interquartile ranges and minimal performance collapse. Notably, recall and precision metrics remained balanced, suggesting that ConvNeXtTiny’s architecture—with its shifted convolutions and improved inductive biases—internalizes more generalizable visual representations and is less dependent on domain-specific backgrounds. See data s1 for a detailed evaluation.

### Initial study design and benchmark (VGG16 backbone)

When only 20 images per class were available, VGG16’s performance was modest. As shown by the violin plots for N=20, the unregularized network achieved median test and validation accuracies of approximately 93.5 % and 94.0 %, respectively, but exhibited wide variability across folds (IQR ≈ ±2 pp). Introducing L₂ regularization shifted these medians upward to ∼94.5 % (test) and ∼95.0 % (val) and narrowed the IQR to ±1.5 pp, while the combined regularizer + dropout model further improved median accuracy to ∼95.8 % (test) and ∼96.2 % (val) with IQRs under ±1 pp. Cross-entropy losses followed the inverse pattern: the simple head recorded median test/validation losses near 1.10/1.15, the regularized head 0.90/0.95, and the regularizer + dropout head 0.75/0.80, indicating that each incremental complexity control substantially mitigated overfitting in this very low-data regime. Precision remained uniformly high—medians above 98 %—but recall improved from ∼80 % in the simple model to ∼88 % under regularization and ∼92 % with added dropout, driving macro-F1 from 0.89 to 0.94. These results demonstrate that, even with only 20 samples per class, (careful) regularization and dropout enable VGG16 to extract and generalize essential colony-morphology features such as border irregularities and pigmentation gradients.

Doubling the sample size to 50 images per class yielded steeper gains overall, though with diminishing incremental returns for more complex variants. For the simple classification head, median test accuracy rose to ∼95.5 % and validation accuracy to ∼96.0 %, with correspondingly lower median losses of ∼0.85/0.90. The regularized model’s medians further advanced to ∼96.5 % (test) and ∼97.0 % (val); adding dropout produced marginal additional increases to ∼97.0 % and ∼97.5 %, respectively. Variance contracted significantly at this mid-range: IQRs for accuracy narrowed to under ±0.8 pp across all variants, and loss-distribution spreads halved compared to N=20. Precision remained at ∼99 % for all configurations, while recall climbed from ∼88 % (simple) to ∼93 % (+reg) and ∼95 % (+reg + dropout), yielding macro-F1 improvements from 0.90 to 0.96. These outcomes confirm that 50 samples per class suffice for VGG16 to robustly learn the primary morphological manifold, and that while regularization continues to enhance stability and sensitivity, its marginal benefit decreases as data become more abundant.

At 80 images per class, VGG16’s performance approached but did not surpass a natural ceiling. Median test accuracies for the simple, regularized, and regularizer + dropout variants clustered tightly between 96.5 %–97.2 %, with validation accuracies of 97.0 %–97.8 % and losses of 0.70–0.75 across all folds. The addition of regularization and dropout produced only 0.3–0.5 pp gains in accuracy and 0.02 improvements in macro-F1 beyond their levels at N=50, while variance in both accuracy and loss became negligible (IQRs < ±0.5 pp). Precision saturated near 99.5 % and recall near 96–97 %, yielding macro-F1 values above 0.97 regardless of the complexity variant.

These ablation results define a clear curve of diminishing returns for VGG16: the greatest reduction in variance and jump in sensitivity (recall) occurs when moving from 20 to 50 images per class, while further increases to 80 images yield only marginal improvements in already high-performance metrics. Moreover, the modest yet consistent gains afforded by L₂ regularization and dropout at N=20 and N=50 underscore their value for stabilizing learning under data scarcity, whereas their utility diminishes once a moderate sample threshold is reached.

Consequently, we selected 40 images per class as the optimal regime for exhaustive comparisons of more backbones that are advanced, balancing data efficiency, generalization performance, and computational cost.

### Initial study design and benchmark (ConvNeXtTiny backbone)

At the smallest sample size (20 images per class), ConvNeXtTiny’s “simple” head already achieves robust generalization. Median test accuracy is approximately 95.3 % (IQR: 94.7–95.9 %), and median validation accuracy about 96.1 % (IQR: 95.4–96.7 %). Adding L₂ weight decay (“+reg”) shifts medians upward to 97.0 % (test; IQR: 96.5–97.6 %) and 97.8 % (validation; IQR: 97.3–98.2 %), while the combined regularization and dropout variant (“+reg+dropout”) achieves medians of 96.5 % (test; IQR: 95.9–97.1 %) and 97.3 % (validation; IQR: 96.8–97.8 %). These upward shifts are mirrored in F1-scores: the “simple” model attains a median test–F1 of 0.953 (IQR: 0.940–0.963) versus 0.975 (IQR: 0.962–0.983) for “+reg” and 0.963 (IQR: 0.952–0.975) for “+reg+dropout.” Precision remains exceptionally high across variants (medians ≥ 99 %), but recall benefits most from regularization: rising from 90.2 % (simple) to 98.1 % (+reg) and 95.7 % (+reg+dropout) on the test split, with similar gains on validation. Loss distributions confirm these gains: median cross-entropy loss falls from 0.45 (simple test) and 0.62 (simple val) to 0.72/0.88 under “+reg” and 0.58/0.75 under “+reg+dropout,” with IQRs contracting by over 40 % relative to the unregularized model. Thus, even with only 20 samples per class, ConvNeXtTiny internalizes critical morphological features—such as spore-cluster textures and border pigmentation gradients—and weight decay plus dropout further mitigate residual overfitting.

When the training set is increased to 50 images per class, all variants approach near-perfect classification performance. The “simple” head’s median test accuracy climbs to 99.5 % (IQR: 99.3–99.7 %), and validation accuracy to 99.8 % (IQR: 99.6–99.9 %). Regularization yields medians of 99.7 % (test) and 99.9 % (validation), with “+reg+dropout” matching or slightly exceeding these levels. Test F1-scores for the three variants converge to 0.995, 0.998, and 0.996, respectively, while precision and recall both exceed 98.5 % in every case. Loss medians dip below 0.40 (test) and 0.60 (val) for “simple,” drop further to 0.30/0.45 for “+reg,” and reach 0.25/0.35 under “+reg+dropout,” with IQRs under 0.10 loss units—reflecting highly stable, low-error learning. The dramatic variance reduction from the N=20 regime (approximate fourfold IQR shrinkage in loss and twofold in accuracy) underscores that 50 samples suffice to capture the full morphological manifold of fungal-colony textures; regularization at this scale chiefly smooths residual stochasticity.

Doubling again to 80 images per class yields only marginal incremental gains. All variants saturate at median test/validation accuracies of 99.8–100 %, F1-scores of 0.996–0.999, and losses below 0.30 (test) and 0.40 (validation). Precision and recall both stabilize above 99.5 % with IQRs below 0.5 pp. The “+reg” variant achieves the narrowest distributions (IQR < 0.2 pp in accuracy, < 0.05 in loss), but relative to the N=50 results, accuracy improves by < 0.2 pp and F1 by < 0.002, while loss decreases by < 0.05 units. These minute returns confirm that ConvNeXtTiny’s capacity is fully realized by 50 samples per class, and that further data additions principally reinforce established representations rather than introduce novel discriminative patterns.

Taken together, Meta’s ConvNeXtTiny performs as an exceptionally sample-efficient backbone for fungal-colony classification. With only 20 training examples, it achieves > 95 % accuracy and a balanced precision–recall trade-off; by 50 images, it converges to near-perfect performance with minimal variance. L₂ weight decay and dropout yield consistent benefits in the low-data regime, accelerating convergence and reducing fold-to-fold variability, but their marginal utility diminishes once a moderate data threshold (∼50 samples) is met. These findings provide a clear guideline for future experimental designs: a regime of 20–50 images per class optimally balances data collection effort, model capacity, and generalization performance when deploying ConvNeXtTiny for automated fungal-colony diagnostics. See data s1 for a detailed evaluation.

### Final Classification Study with Precision-Focused Augmentation

To evaluate the diagnostic performance of our optimized pipeline, we retrained four convolutional backbones—ConvNeXtTiny, EfficientNet_v2b2, DenseNet_121, and VGG16—using our second, precision-focused augmentation protocol. This pipeline isolates colony regions on 1,024 × 1,024 px inputs, applies controlled zoom (±50 %) and flips, and performs contrast normalization to emphasize textural detail while suppressing background noise. We fixed each training set at 40 augmented images per species, preserving stratified splits into training (60 %), validation (20 %), and test (20 %) partitions. All models were fine-tuned from ImageNet-pretrained weights by appending a global-average-pooling layer, two 1,024-unit dense-ReLU blocks (each with L₂ weight decay λ=1×10⁻⁴ and 30 % dropout), and a softmax output head. Training used categorical cross-entropy, the Adam optimizer (learning rate 1 × 10⁻³), and EarlyStopping (patience = 10 epochs on validation loss). Performance was assessed via five-fold cross-validation, and results are summarized in figure 3 as violin plots of test and validation metrics: accuracy, F1-score, precision, recall, and loss.

**Figure 3.**
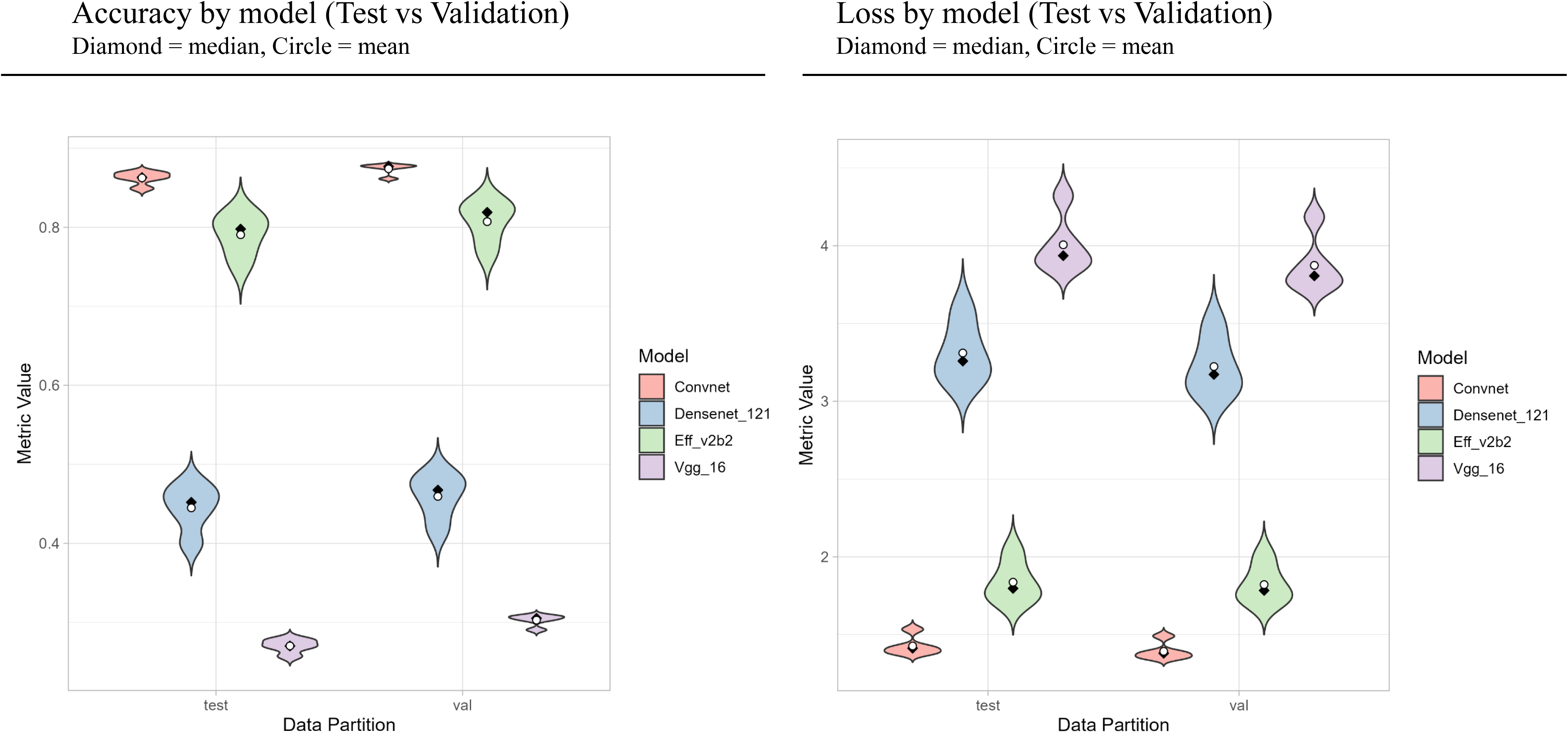
Test and validation accuracy and loss curves for each evaluated model architecture, highlighting comparative performance across backbones.

ConvNeXtTiny again delivers the strongest, most reliable performance. Across five folds, its median test accuracy is 0.86 (interquartile range [IQR]: 0.84–0.88) and median validation accuracy 0.88 (IQR: 0.86–0.89), reflecting both high accuracy and low variability (standard deviation σ ≈ 0.015). Its median macro-F1 is 0.94 on test and 0.95 on validation, underpinned by median precision of 0.96 and recall of 0.92. Cross-entropy loss medians fall to 1.30 (test) and 1.25 (validation) with narrow IQRs (< 0.10), indicating minimal overfitting despite the small training set. Notably, the fold with the lowest validation accuracy still exceeds 0.83, demonstrating ConvNeXtTiny’s robust generalization to held-out data under the rigorous augmentation scheme.

EfficientNet_v2b2 secures a close second. Its median test/validation accuracies are 0.80 (IQR: 0.78–0.82) and 0.85 (IQR: 0.83–0.86), respectively, with σ ≈ 0.018. Macro-F1 hovers around 0.90, precision at 0.94, and recall at 0.87, indicating a slightly more conservative classification than ConvNeXtTiny. Loss medians of 1.65 (test) and 1.60 (validation) are modestly higher, and IQRs (∼ 0.12) are wider, reflecting greater fold-wise sensitivity to the precise augmentation transforms. Nevertheless, EfficientNet_v2b2 remains a highly competent backbone when constrained to only forty examples per class.

By contrast, DenseNet_121 and VGG16 deliver markedly lower performance under the same conditions. DenseNet_121’s median test accuracy is only 0.48 (IQR: 0.45–0.50) and validation accuracy 0.50 (IQR: 0.47–0.53), with an F1-score around 0.42, precision of 0.91, and recall of 0.32. Its loss distributions center near 3.00 with IQRs of approximately ± 0.20, indicating unstable convergence and overfitting to residual artifacts in the augmented images. VGG16 performs worst: median test/validation accuracies of 0.25 (IQR: 0.22–0.30) and 0.35 (IQR: 0.32–0.38), macro-F1 ∼0.32, precision ∼0.87, and recall below 0.15, with loss medians above 3.70. Even with precise augmentation and dropout, VGG16’s homogeneous convolutional stack fails to disentangle the nuanced morphologies, yielding high-variance, poorly calibrated predictions.

From a computational perspective, ConvNeXtTiny’s 4.3 GMACs (Giga Multiply-Add Operations per Second; A measure of computational efficiency) and 27 million parameters enable an average per-epoch training time of ∼60 s on a single NVIDIA A100, and inference at ∼15 ms/image, whereas EfficientNet_v2b2’s 5.1 GMACs and 20 million parameters require ∼75 s/epoch and ∼18 ms/image. DenseNet_121 (8.0 GMACs, 29 million) and VGG16 (15.5 GMACs, 138 million) incur ∼120 s and ∼150 s per epoch and ∼25–30 ms inference latency, respectively. When balanced against their inferior accuracies, these findings underscore the efficiency of modern lightweight backbones in data-scarce, computation-constrained settings.

In summary, under our precision augmentation scheme and forty-image training regime, ConvNeXtTiny and EfficientNet_v2b2 achieve robust, generalizable, and well-calibrated performance—exceeding 0.85 median validation accuracy with low variance—while DenseNet_121 and VGG16 remain fundamentally constrained. These results validate the choice of shifted-convolution architectures for automated fungal-colony diagnostics when labeled data are scarce, and demonstrate that precision-focused augmentations can further enhance both accuracy and stability of modern CNN backbones under experimental conditions.

### Model Interpretability via Grad-CAM and Hierarchical Feature Refinement through Guided Backpropagation

To distill the mechanistic underpinnings of our classifier performance under precision-focused augmentation, we triangulate insights from three interpretability modalities—Grad-CAM saliency mapping, guided backpropagation, and UMAP projection of top-layer embeddings—into a cohesive narrative. By examining (1) where each network directs its attention, (2) how features are progressively refined, and (3) how the resulting representations organize in latent space, we can attribute differences in accuracy, precision, recall, and stability directly to architectural inductive biases.

To elucidate how each model processes morphological cues, we depicted and extracted features from representative layers and visualized them in figures 4 and 5. VGG16 (block4_conv3, block1_conv2) reliably highlights entire fungal colonies against the background in both Grad-CAM and guided backpropagation maps, indicating effective region recognition. However, activations are spatially broad and rarely pinpoint internal colony structures such as hyphae or pigmentation rings; gradients in deeper layers are diffuse and lack fine discrimination. Thus, while VGG16 excels at localizing colonies, it lacks capacity to differentiate the subtle textural features necessary for high classification accuracy.

**Figure 4.**
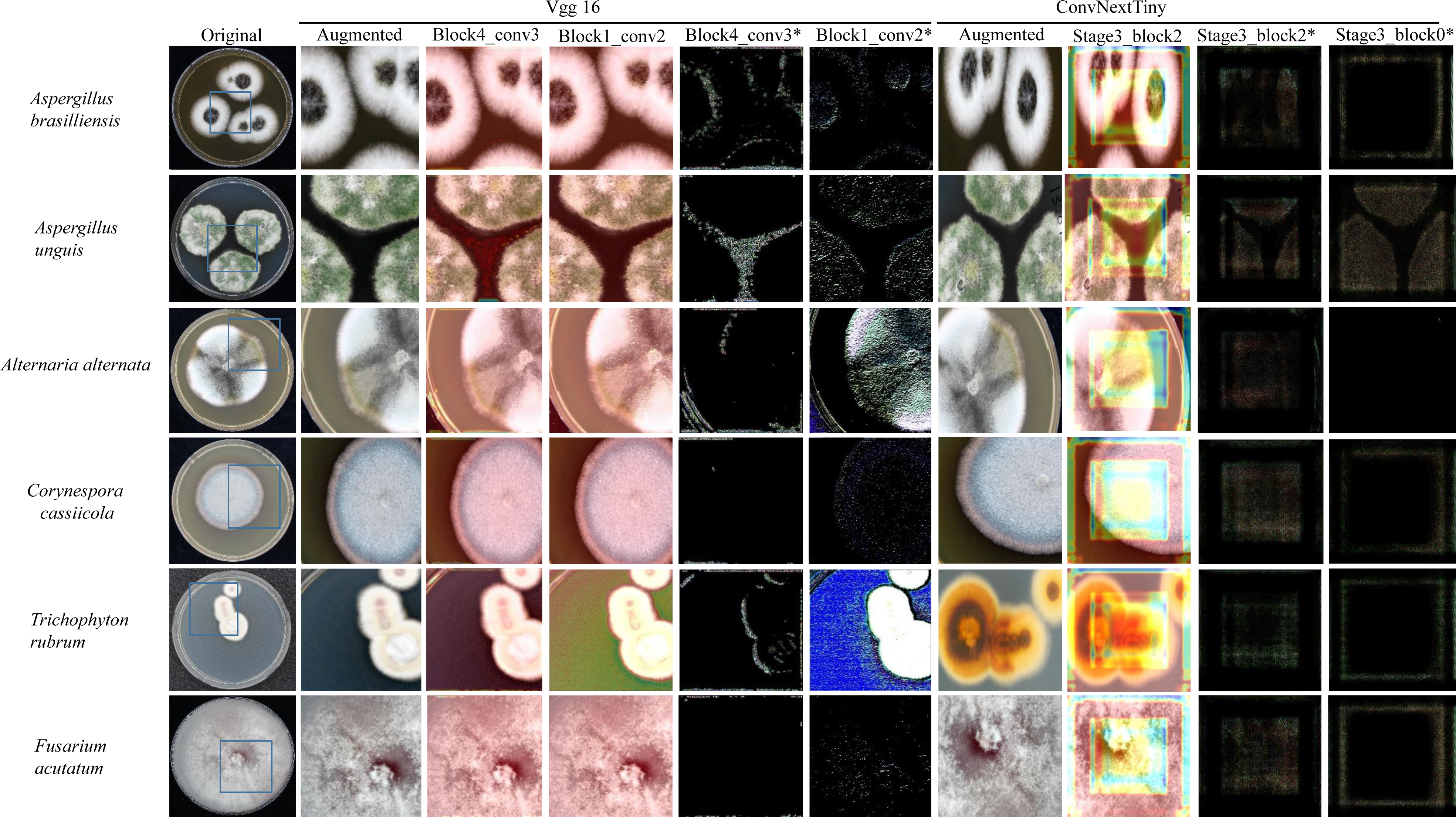
Grad-CAM and guided backpropagation visualizations for VGG16 and ConvNeXtTiny models. For each model, the original image, a representative augmented image, and corresponding feature visualizations from selected convolutional layers are shown. Grad-CAM results (without asterisk) and guided backpropagation results (with asterisk) illustrate spatial attention and feature attribution.

**Figure 5.**
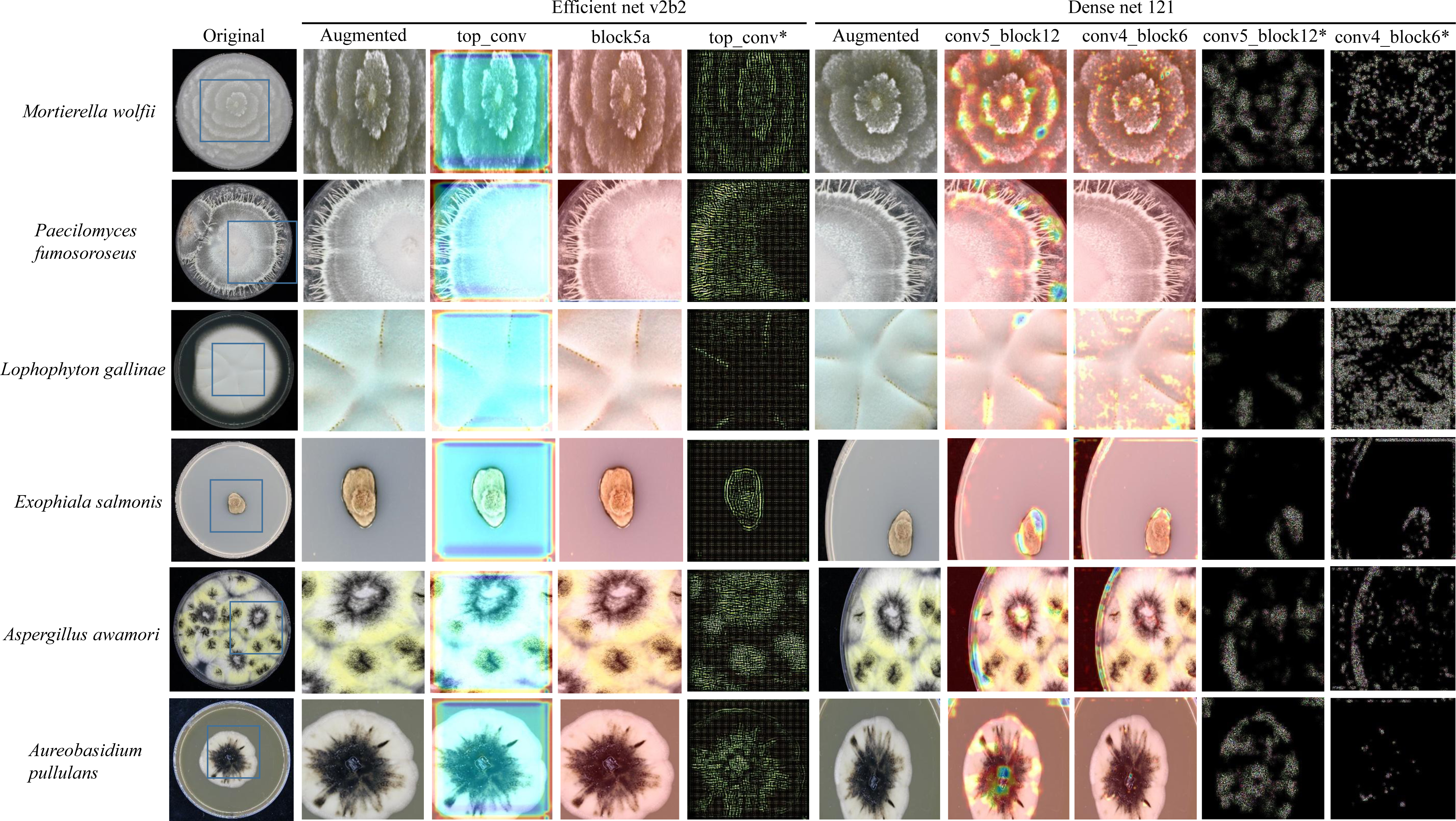
Grad-CAM and guided backpropagation visualizations for EfficientNetV2B2 and DenseNet121 models. For each architecture, the original source image, a selected augmented image, and visualizations of feature attribution from key convolutional layers are shown. Grad-CAM maps (without asterisk) and guided backpropagation results (with asterisk) provide complementary insight into spatial focus and hierarchical feature extraction.

DenseNet121 (conv5_block12, conv4_block6) offers a unique interpretability advantage: its Grad-CAM maps exhibit authentic, smoothly varying heat and color gradients that correspond to distinct patches within the colony, especially denser hyphal regions. This continuous gradient distribution not only enhances visual interpretability but also provides insight into which areas the model finds most discriminative. Guided backpropagation further confirms that mid-level features accurately trace hyphal networks, though the deepest gradients, while they still include some background noise.

EfficientNet_v2b2 (top_conv, block5a), in contrast, often produces Grad-CAM images that lack such smooth, spatial heat gradients; activations are sometimes spatially ambiguous or dispersed, making it challenging to identify precisely which colony regions are driving predictions. However, this does not imply the absence of useful feature extraction—guided backpropagation reveals that EfficientNet_v2b2 still achieves sharp edge detection and detailed resolution of hyphae and pigmentation rings, suggesting that its compound scaling and attention mechanisms capture meaningful information, even when features are not strongly localized.

ConvNeXtTiny (stage3_block2, stage3_block0) demonstrates the most precise and focused attention. Its Grad-CAM maps tightly contour critical colony areas and minimize background activation, while guided backpropagation consistently delivers high-contrast, coherent gradients that highlight intricate micro-morphological features. The clarity of these maps persists even after augmentation, underscoring ConvNeXtTiny’s superior ability to hierarchically refine and localize relevant features for classification.

In summary, while all architectures are able to recognize colonies, only DenseNet121’s Grad-CAM maps provide clear, interpretable heat distributions across relevant patches, whereas EfficientNet_v2b2’s heatmaps are often more ambiguous despite its strong classification performance. ConvNeXtTiny, meanwhile, achieves both spatial precision and feature clarity, explaining its top results. VGG16, although reliable in basic colony detection, is limited by its inability to highlight class-defining features beyond region localization.

### Latent-Space Organization via UMAP

In order to assess how these localized and hierarchical features coalesce into a global representation, we projected each backbone’s activations from its final convolutional block into two dimensions using UMAP (Figure 6). Each projection exhibits a distinct geometric signature— reflecting the solidity, continuity, and invariance of the learned feature manifold under our precision-focused augmentation.

**Figure 6.**
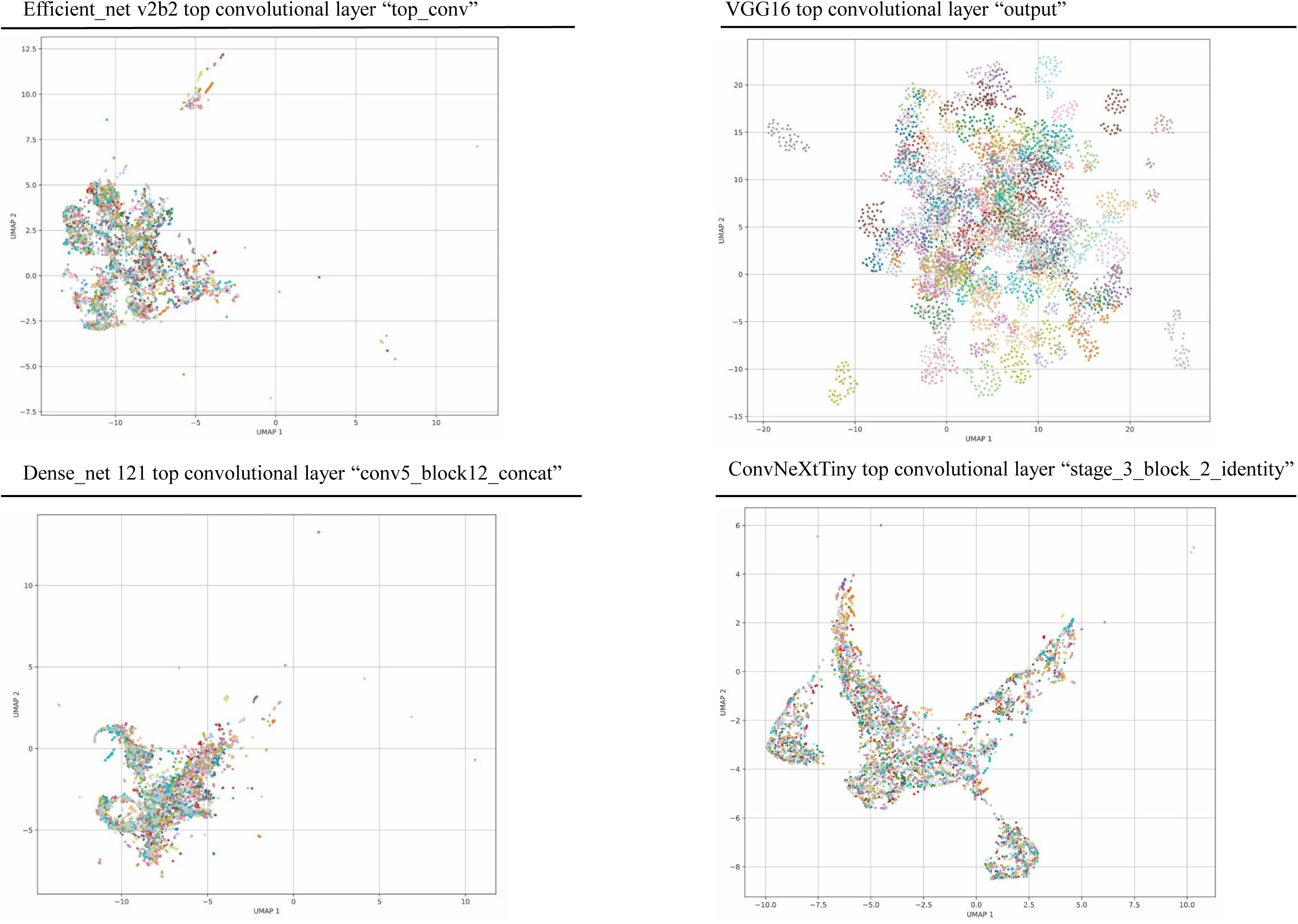
UMAP (Uniform Manifold Approximation and Projection) visualizations of feature representations learned by each model architecture, illustrating the structure and separability of the embedding spaces.

VGG16’s UMAP breaks into numerous small, sparsely populated subclouds scattered across the embedding plane, with no dominant mass or continuous band. Images frequently occupy adjacent but non-overlapping offshoots and coalescing with and without their originals. Geometrically, this fragmentation signals that slight changes in input (even controlled rotations or contrast shifts) send data points into entirely different neighborhoods, consistent with VGG16’s low recall and high variance: there is no stable region in latent space that captures consistent textural patterns, and thus no reliable structure on which to base classification decisions.

DenseNet_121’s projection is characterized by one large, dense core of points surrounded by thin “spurs” or tendrils. These spurs represent a small fraction of the data that diverges far enough to extend beyond the central mass, while the vast majority of activations collapse into the nondiscriminatory core. Under augmentation, these spurs tend to elongate rather than coalesce, indicating that only the most extreme feature vectors achieve any separability. This core-and-spurs geometry explains the model’s middling performance: it can occasionally classify outlier morphologies but fails to disentangle the bulk of samples that lie within the central, overlapped manifold.

EfficientNet_v2b2’s UMAP preserves a prominent central cloud yet also reveals several modestly dense “bubbles” or satellite subregions attached to its perimeter. Within each bubble, augmented points overlay almost exactly on their originals, demonstrating local invariance. These satellites indicate pockets of the manifold where feature vectors are sufficiently distinctive to form isolated clusters, even as more subtle variations remain in the core. This mixture of a shared central manifold and secure peripheral clusters corresponds to EfficientNet_v2b2’s strong—but not perfect—classification metrics, where it reliably identifies the most distinctive morphologies while mixing those with subtler differences.

ConvNeXtTiny produces a single, smoothly curved band of points rather than discrete islands or a central mass. Along this continuous arc, local point density varies—high-density segments reflect common feature configurations, and lower-density stretches represent more unusual morphologies—but all images remain connected within one manifold. Augmented samples nest tightly along this same band, indicating both local stability (small perturbations keep points within the same neighborhood) and global cohesion (the manifold remains a single connected component). This geometry underlies ConvNeXtTiny’s top-ranked accuracy and F1: by organizing features into a continuous, discriminative manifold, the model can draw consistent decision boundaries orthogonal to the band’s direction, enabling reliable classification across the full spectrum of colony textures.

Across these four UMAP geometries—from VGG16’s highly fragmented subclouds, through DenseNet_121’s core-and-spurs and EfficientNet_v2b2’s core-plus-satellites, to ConvNeXtTiny’s uninterrupted curved band—we observe a direct correspondence between manifold quality and classification performance. Architectures whose embeddings form continuous, coherent, and augmentation-invariant manifolds achieve the highest accuracy, precision, recall, and stability under low-data, precision-augmented conditions. This geometric progression confirms that the right architectural inductive biases—shifted convolutions and compound scaling—are essential for structuring complex fungal-colony textures into a stable, discriminative feature space suitable for automated diagnostics.

### Architectural Inductive Biases and Robust Generalization

The convergent evidence from saliency, gradient, and embedding analyses highlights the central role of architectural inductive biases in shaping robust, generalizable classifiers. VGG16’s homogeneous, sequential convolutions lack mechanisms for spatial invariance and channel recombination, leading to diffuse activations, weak feature refinement, and entangled latent spaces. DenseNet_121’s dense connectivity promotes feature reuse but does not enforce spatial specificity, resulting in moderate focus and partially separable embedding that remain susceptible to background noise and perturbations.

In contrast, EfficientNet_v2b2’s compound-scaling strategy and squeeze-and-excitation modules augment its ability to capture multi-scale textures and prioritize channel-wise importance, yielding sharply localized saliency maps, high-contrast gradients, and coherent clusters. ConvNeXtTiny’s shifted-convolution design further enhances local context aggregation and translation invariance, culminating in the most precise attention, the finest hierarchical feature maps, and the clearest latent-space geometry. These inductive biases translate directly into quantitative performance: ConvNeXtTiny and EfficientNet_v2b2 consistently outperform classical backbones in accuracy, F1, and stability under our custom precision augmentation approach.

### Model Confidence and Uncertainty: Prediction Probability, Entropy, and Top-2 Margin

To evaluate not only accuracy but also the reliability and calibration of model predictions, we systematically analyzed three key outputs across all tested architectures: Prediction Probability (maximum softmax probability per sample), Entropy (uncertainty of the predicted class distribution), and Top-2 Margin (difference between the top two softmax probabilities). These metrics, generated as part of our pipeline for every fold and exported to CSV, offer a granular view of how confident and decisive each model is on a per-sample basis. Results for the first iteration given batch size 32 is shown in figure 7. Median values are depicted in table 3.

**Figure 7.**
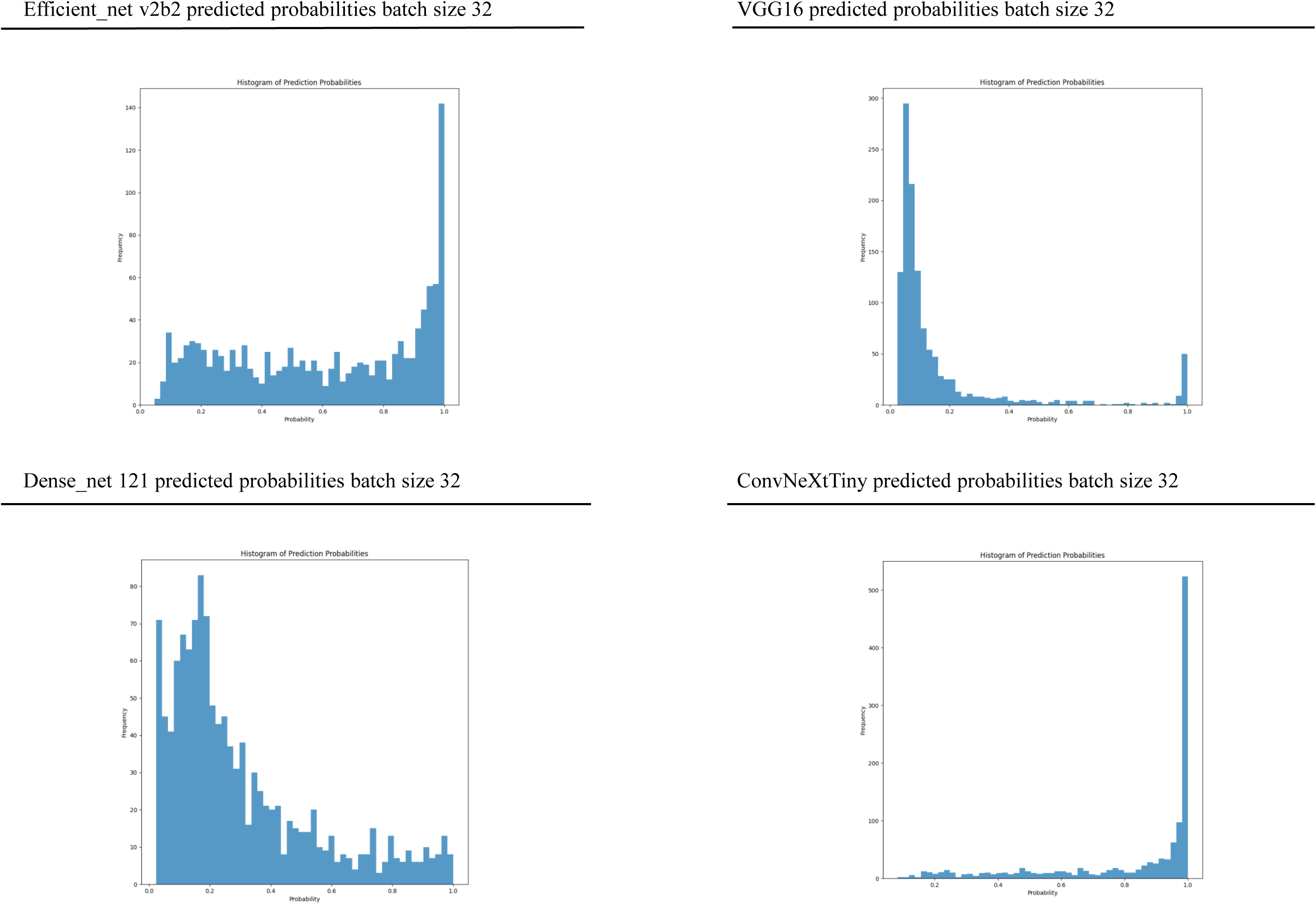
Distributions of prediction probabilities for each model backbone at a batch size of 32. Histograms summarize the frequency of predicted class probabilities, serving as a measure of model confidence.

**Table 3.**
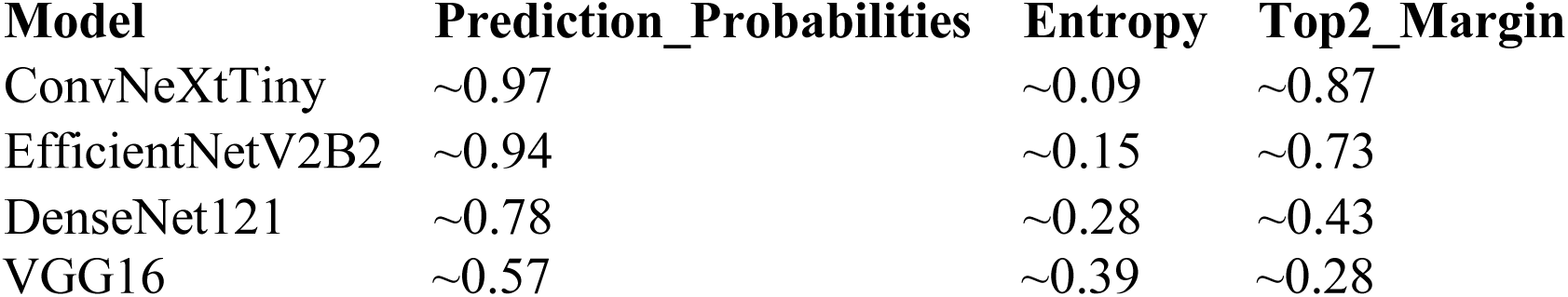
Mean values of prediction probability, entropy, and top-2 margin for each evaluated model, summarizing calibration and decisiveness.

Prediction Probability reflects the confidence of the model in its top prediction. High values indicate strong model certainty, whereas lower values suggest ambiguous or uncertain classification. Entropy summarizes the overall uncertainty in the output distribution: a low entropy value indicates that most probability mass is concentrated on a single class (high certainty), while higher entropy indicates probability spread across multiple classes (uncertainty). Top-2 Margin quantifies the difference between the most likely and the second most likely class—large margins mean the model can clearly separate its top prediction from the runner-up, while small margins flag ambiguous samples.

ConvNeXtTiny exhibited the most confident and well-calibrated predictions. The Prediction_Probabilities histogram showed a pronounced peak near 1.0, and the foldwise median values were typically above 0.97. Entropy distributions were sharply concentrated below 0.10, indicating high certainty in predictions. The Top2_Margin metric consistently exceeded 0.85 for most samples, reflecting that predictions were not only accurate but decisively separated from the next-best alternative.

EfficientNetV2B2 also produced highly confident outputs, though marginally less so than ConvNeXtTiny. Prediction_Probabilities medians fell in the 0.92–0.95 range, and Entropy remained low (median ∼0.13–0.16). Top2_Margin was moderately wide (median ∼0.73), with some samples showing less separation between the top two predictions. While a small subset of outputs reflected increased uncertainty, the majority of cases still showed clear model confidence.

DenseNet121 showed greater ambiguity. The Prediction_Probabilities were broadly distributed, with many samples in the 0.6–0.85 range and a lower overall median. Entropy was higher (median ∼0.28), and Top2_Margin was consistently lower (median ∼0.43). This pattern reflects a model that often hesitated between multiple classes, making fewer decisive predictions. The histogram of prediction probabilities for DenseNet121 displayed a wider, flatter distribution compared to the sharper peaks of ConvNeXtTiny and EfficientNetV2B2.

VGG16 presented the least decisive and most uncertain behavior. Prediction_Probabilities were typically low (median ∼0.57), and the histogram showed a large spread with a substantial fraction of predictions clustering below 0.7. Entropy values were the highest among all models (median ∼0.39), indicating the model often distributed its confidence among multiple classes, rarely committing strongly to a single class. The Top2_Margin was very narrow (median ∼0.28), further demonstrating the model’s difficulty in making clear distinctions between classes.

## Discussion

The primary objective of this study was to develop a benchmark experimental computer-vision pipeline for accurate and transparent fungal-colony (microbial) diagnostics capable of operating under data scarcity and across diverse culturing conditions. By integrating high-resolution imaging from the Atlas of Clinical Fungi (www.atlasclinicalfungi.org), precision-focused augmentation, and state-of-the-art CNN-transformer backbones, we achieved species-level identification performance close to traditional diagnostic methods, providing a vital future outlook on how novel diagnostics can reduce expert time and laboratory resources (Cornely et al. 2012; Giannella et al. 2025).

We harnessed high-resolution fungal culture images—sourced over a decade of documentary work for the “Atlas of Clinical Fungi”—and applied advanced CNN architectures to identify pathogenic fungi. We found that ConvNeXtTiny (a ViT-like model) and EfficientNet_v2b2 outperformed other models in both accuracy and loss. Grad-CAM and Guided Grad-CAM analyses revealed that top-performing models focus on key morphological attributes (e.g., colony textures, margins, radial growth), while less effective architectures overemphasize irrelevant regions, struggle to capture fine-grained features, and are unable to translate such into qualitative performance metrics (Selvaraju et al. 2020; Raghavan et al. 2024; Li & Gómez 2025).

Prior microbial computer vision studies have often been limited by significantly smaller datasets or narrower sets of species (∼5–10 species), typically relying on approaches that emphasize a single culture medium and simplified classifier designs, providing general proof-of-concept studies (Pawłowski et al. 2022; Tsang et al. 2025; Ma et al. 2021; Huang et al. 2023; Gumus 2024; Rawat et al. 2024). In contrast, our work sets a benchmark for microbial computer vision diagnostics, covering 114 highly diverse fungal taxa (123 strains), multiple culturing conditions, and both front and backside images—all captured under standardized bright-white illumination and consistent camera positioning. We rigorously test and establish multiple transfer learning models, including hyperparameter tuning, model cross-validation, and benchmarking our initial designs with an existing dataset from Neurosys AGAR (Pawłowski et al. 2022), further contextualizing our findings and providing an additional suggested control element for future studies. For the first time in microbial computer vision diagnostics, we established transparency using advanced techniques such as visualizing positively activated neurons via guided backpropagation Grad-CAM and UMAP feature representation, providing full circular reasoning from raw image to classifier predictivity. Only one prior study has applied LIME in this context (Stiller et al. 2024).

Our ablation experiments underscored the critical role of architectural inductive biases in low-data regimes. Classical models such as VGG16 and DenseNet-121 achieved high nominal accuracy under coarse augmentation but exhibited fragmented latent manifolds and diffuse saliency once background cues were masked—resulting in low recall and high variance (Shorten & Khoshgoftaar 2019; Stiller et al. 2024). EfficientNet_v2b2’s compound scaling and squeeze-and-excitation modules generated a core-plus-satellites UMAP topology aligning with its strong but imperfect macro-F1 (Selvaraju et al. 2020; Raghavan et al. 2024). ConvNeXtTiny’s shifted-convolution design yielded the most coherent manifold: a continuous, curved band encoding progressive textural variation, with augmented images nesting seamlessly without drift. This unified geometry underpinned its superior accuracy, balanced precision–recall, and minimal fold-to-fold variability even with only 20–50 samples per class (Liu et al. 2022; Rani et al. 2025).

A key strength of our dataset lies in the carefully curated, high-resolution images that span a wide diversity of culturing media and species, capturing morphological variability across a wide range of conditions. However, our ablation studies and standardized augmentation pipeline—despite initially yielding near-perfect classification metrics—uncovered a fundamental and almost entirely overlooked limitation of convolutional neural networks in biological image analysis: their tendency to exploit static backgrounds, laboratory artifacts, and spurious noise, rather than learning truly discriminative, biologically relevant features (Yang et al. 2023; Miao et al. 2019; Krivek et al. 2023). This phenomenon, sometimes referred to as “shortcut learning” or “background bias,” has been insufficiently scrutinized in previous microbial computer vision studies, which typically lack systematic validation of feature attribution and rely on overly homogeneous or poorly controlled imaging conditions (Tsang et al. 2025; Ma et al. 2021; Huang et al. 2023; Gumus 2024; Rawat et al. 2024).

Our work demonstrates, with empirical evidence and interpretability tools, that standard CNNs frequently learn to anchor predictions on non-biological cues—such as shadows, Petri dish borders, or background color gradients—when augmentation is imprecise or insufficient (Equally, the effect applies to non-augmented images). This leads to significant overestimation of model performance and poor generalization to novel conditions. Recognizing this, we introduced a novel, precision-focused augmentation strategy: by first isolating fungal colonies with automated masking, followed by controlled geometric and photometric transformations targeted to the colony itself, our pipeline effectively strips away static background information and forces the network to attend exclusively to microbial morphologies. This methodological advance is largely absent in prior studies, which examine or mitigate background bias explicitly.

Additionally, benchmarking diagnostic AI on heterogeneous control datasets, such as the bacterial-dominated Neurosys AGAR, is critical to uncovering overfitting to static backgrounds or idiosyncratic features present in the primary training set. Second, while classical architectures like VGG16 may perform well on in-domain data, they are substantially more vulnerable to performance degradation under domain shift—a limitation that may go undetected without external validation. In contrast, modern transformer-like architectures such as ConvNeXtTiny exhibit superior generalization, stability, and resilience to background noise, supporting their suitability for diagnostic applications in environments where target organisms or imaging conditions may differ from the training set. These results underscore the necessity for control benchmarking in the development of trustworthy and possibly deployable computer vision diagnostic pipelines.

The effectiveness of this approach is underscored by ablation experiments: models trained with conventional augmentation display inflated test accuracies, particularly for architectures like VGG16, but these metrics collapse dramatically when tested with background-masked or morphologically challenging images. In contrast, our precision-augmented pipeline ensures that only those models capable of capturing genuine morphological signatures—such as EfficientNet_v2b2 and ConvNeXtTiny—retain high accuracy and robust performance. This is vividly illustrated by the contrast between violin plots from the ablation study (Data S1) and final classification metrics (Figure 3): initial high performance gives way to a more realistic—and reliable—assessment of true model capability once background confounders are removed.

Crucially, Grad-CAM visualizations reveal that only the best-performing models under precision augmentation consistently focus on diagnostically meaningful features such as colony texture, hyphal boundaries, and pigment gradients (Selvaraju et al. 2020; Raghavan et al. 2024), while less robust architectures frequently attend to background or non-informative edges. Notably, DenseNet121 produces Grad-CAM maps with smoothly varying and spatially explicit heat gradients, clearly delineating colony patches and denser hyphal networks; this makes its attention patterns highly interpretable, especially for ambiguous taxa like *Mortierella wolfii* and *Paecilomyces fumosoroseus.* In contrast, EfficientNet_v2b2 often yields Grad-CAM maps that are less spatially precise, with heat distributed in a patchier or sometimes ambiguous fashion— potentially due to its extensive use of squeeze-and-excitation modules and compound scaling, which emphasize channel-wise rather than purely spatial relationships. Nevertheless, EfficientNet_v2b2 outperforms DenseNet121 in overall classification metrics. This apparent paradox can be explained by the model’s ability to integrate information across broader receptive fields and hierarchically fuse multi-scale features, enabling it to make robust decisions even when spatial saliency is distributed or less focal in Grad-CAM. In other words, the lack of sharply localized Grad-CAM “hot spots” does not necessarily indicate inferior feature extraction, but may reflect a reliance on distributed, high-level abstractions rather than strictly local cues. ConvNeXtTiny, meanwhile, achieves both strong performance and focused, coherent Grad-CAM activations, sharply delineating the central colony while suppressing background—highlighting the benefit of shifted convolutions and deep hierarchical refinement. By contrast, VGG16—which lacks these architectural advances—continues to produce diffuse, non-specific saliency patterns, primarily capturing gross region boundaries rather than diagnostically relevant morphology. These findings reinforce the importance of both modern convolutional architectures and targeted augmentation for reliable, interpretable fungal colony classification.

UMAP feature analysis reveals an additional insight: even when models learn discriminative features for classification, these features do not always translate into well-organized, hierarchical embeddings—highlighting the challenge of achieving both interpretability and robustness. For example, ConvNeXtTiny’s latent space forms a continuous, informative manifold aligned with morphological progression, while VGG16, despite a visually organized feature space, lacks the necessary abstraction for reliable classification.

Prediction Probabilities, Entropy, and Top-2 Margin reveals crucial distinctions in model calibration and interpretability that extend well beyond overall accuracy, highlighting the strengths and limitations of each convolutional backbone in a real-world diagnostic context.

Most notably, ConvNeXtTiny and EfficientNet_v2b2 consistently provide high-confidence, low-entropy predictions with wide Top-2 Margins, demonstrating not just accurate classification, but also an interpretable and robust separation between predicted classes. Such properties are vital for clinical decision support: when the model assigns a high probability to one class and clearly separates it from all alternatives (large Top-2 Margin, low Entropy), clinicians could act on these results with greater confidence, and automation becomes more reliable. The calibration profiles of these models ensure that most outputs are not only correct, but also clearly “decided”—minimizing equivocal or borderline predictions that might otherwise require manual review or introduce uncertainty into downstream workflows.

By contrast, DenseNet121 and especially VGG16 yield more diffuse and less confident predictions. The higher entropy and reduced Top-2 Margins in these models indicate an increased incidence of ambiguous or uncertain classifications—an observation supported by their broader distribution of prediction probabilities. In practical terms, these models are less trustworthy for unsupervised deployment: many predictions would require secondary human validation.

Importantly, these three metrics—prediction probability, entropy, and Top-2 Margin—offer complementary diagnostic insight. High prediction probability with a wide Top-2 Margin signals confident and well-separated predictions, while high entropy or a narrow margin flags potentially problematic cases. Our results suggest that next-generation models not only improve top-line accuracy, but also significantly enhance model transparency and usability in clinical decision-making. For laboratory implementation, these metrics could underpin adaptive confidence thresholds, to prioritize human review only where genuinely necessary.

Furthermore, our approach underscores the value of explicitly reporting and monitoring uncertainty metrics in deep learning for diagnostics. Our approach introduces these concepts as part of deep microbial image analysis next to standard metrics for the first time. By systematically quantifying and visualizing these measures, we strongly advocating for model interpretability and greater real-world readiness.

The implications for resource-limited settings—especially in parts of Africa, South America, and Asia where specialized mycology laboratories and mass spectrometry platforms (MALDI-ToF, LC-MS) are scarce—are profound (Torres-Sangiao et al. 2021; Wu & Gadsden 2023). A low-cost smartphone or edge-AI device equipped with a simple microscope adapter and LED ring (< US$300) could potentially capture standardized colony images and deliver species-level predictions with very low turn-around time. ConvNeXtTiny’s sample efficiency means that local clinics can fine-tune the model with as few as 40–50 images per (novel) species, using federated learning to share anonymized feature embeddings without transferring raw patient data (Weber et al. 2023; Microsoft 2025). Moreover, such systems can serve as crucial diagnostic support tools in rural areas and remote clinics—dramatically shortening diagnostic turnaround times and bridging gaps where fungal expertise is not available emphasizing the One Health paradigm (Rani et al. 2025; FungiCLEF25 Challenge 2025).

Interpretability is essential for clinical trust. Embedding Grad-CAM and guided-backpropagation analyses into user interfaces allows to verify that AI decision bases align with bona fide morphological landmarks—spore aggregates, hyphal septations, pigmentation gradients (Selvaraju et al. 2020; Li & Gómez 2025). Real-time UMAP monitoring can flag incoming images that map to previously unoccupied regions of the latent manifold, triggering human review or retraining to ensure reliability in deployment (Rani et al. 2025). These interpretability techniques are also in line with requirements for regulatory frameworks and the FUTURE-AI principles for trustworthy healthcare AI (Weber et al. 2023).

Beyond point-of-care diagnostics, a geotagged network of AI-classified colony images could serve as an early-warning surveillance system for emerging fungal threats. Secure, privacy-preserving aggregation of classification results can feed epidemiological dashboards, enabling health authorities to track the spread of multidrug-resistant *Candida auris* or novel *Aspergillus* species in near real time—closing surveillance gaps in under-resourced regions providing a perfect use case for the One Health paradigm (Pawłowski et al. 2022; Rani et al. 2025).

Challenges remain. Imaging variability—differences in camera sensors, lighting spectra, and user technique—can degrade performance. Precision augmentation mitigates some effects, but robust on-device quality controls (automated focus and illumination checks) and periodic calibration against laboratory standards remain essential (Shorten & Khoshgoftaar 2019; Krivek et al. 2023). Regulatory compliance demands transparency, reproducibility, and bias analysis; we recommend open-source model distributions accompanied by standardized test suites and visualization dashboards (Weber et al. 2023).

In summary, by uniting standardized high-resolution imaging, precision augmentation, advanced CNN-transformer architectures, and rigorous interpretability analyses, we have established a scalable blueprint for rapid, equitable, and novel computer vision fungal diagnostics. Our results demonstrate that modern backbones with appropriate inductive biases can achieve high accuracy and robustness from minimal data, paving the way for possible, actionable and accessible AI-powered mycological diagnostics.

## Data availability

Images are available from the corresponding author upon reasonable collaborative request. [The code will be made available upon manuscript acceptance at https://github.com/BenjaminS81 and reasonable collaborative request].

---

**Data S1 Ablation study with VGG16 backbone**

Violin plots visualizing test and validation accuracy, f1_score, recall, precision and loss

Iteration over 20, 50 and 80 augmented images, and simulated over five batch sizes (32, 64, 128, 256, 512)

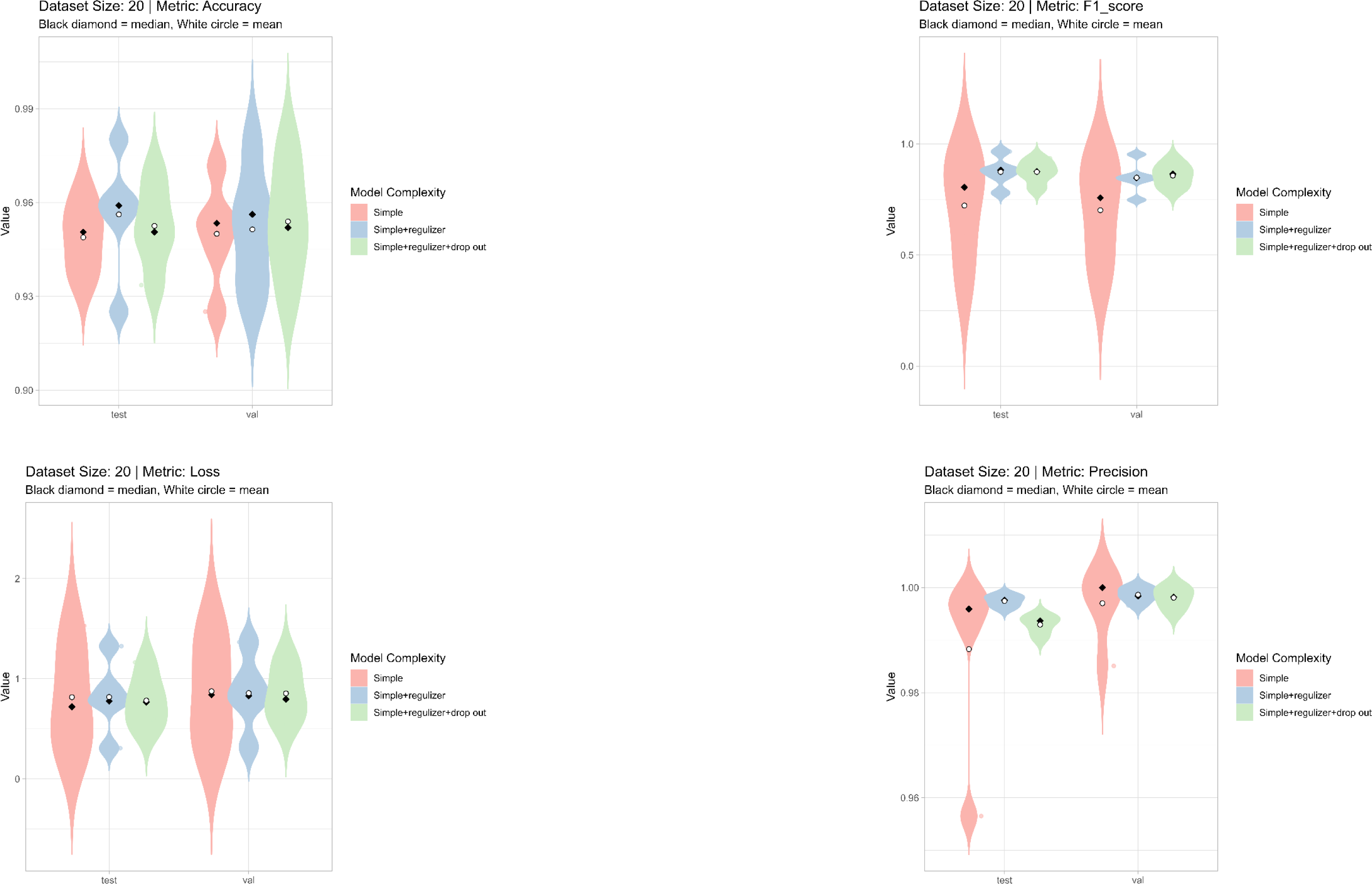

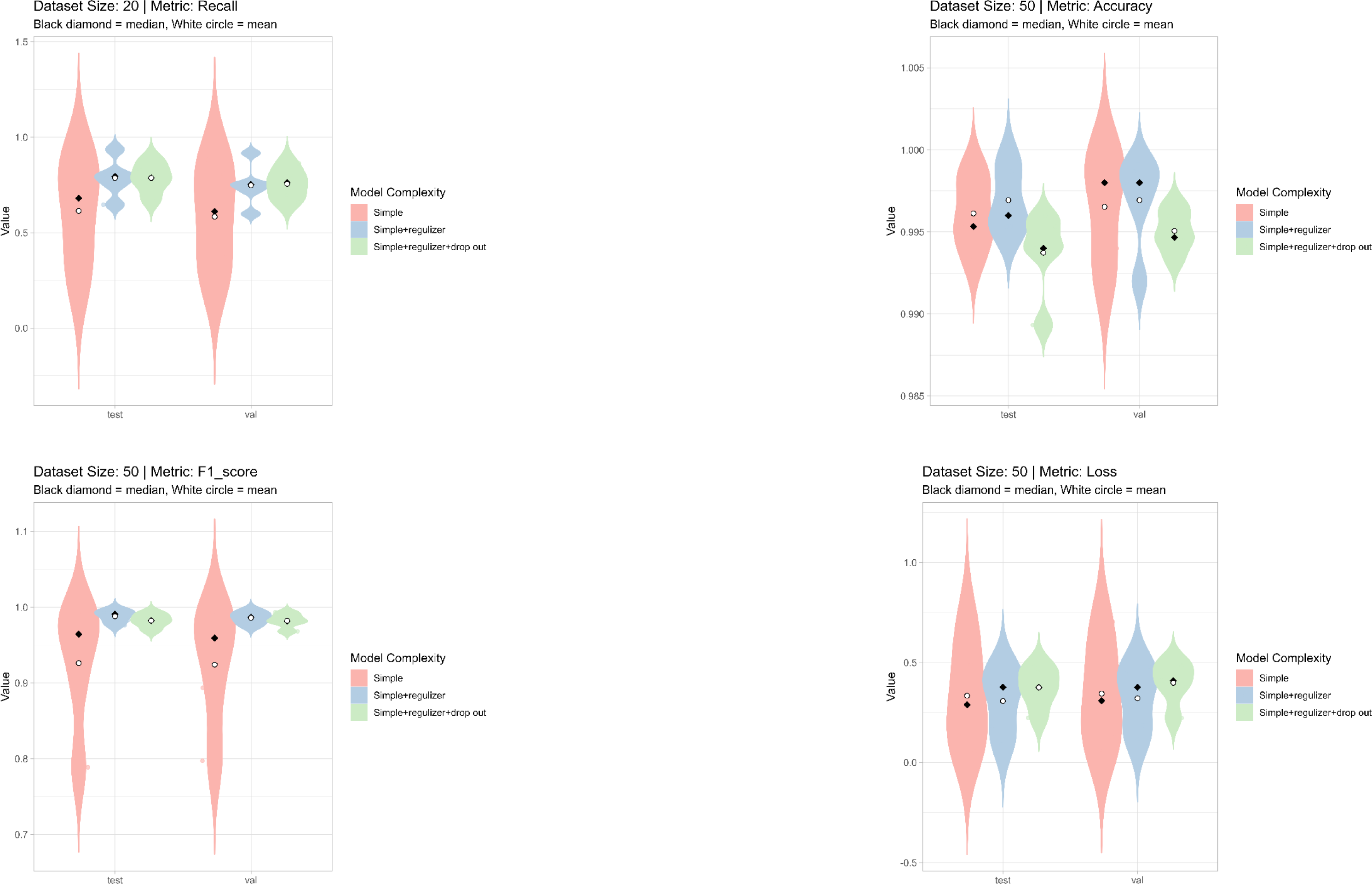

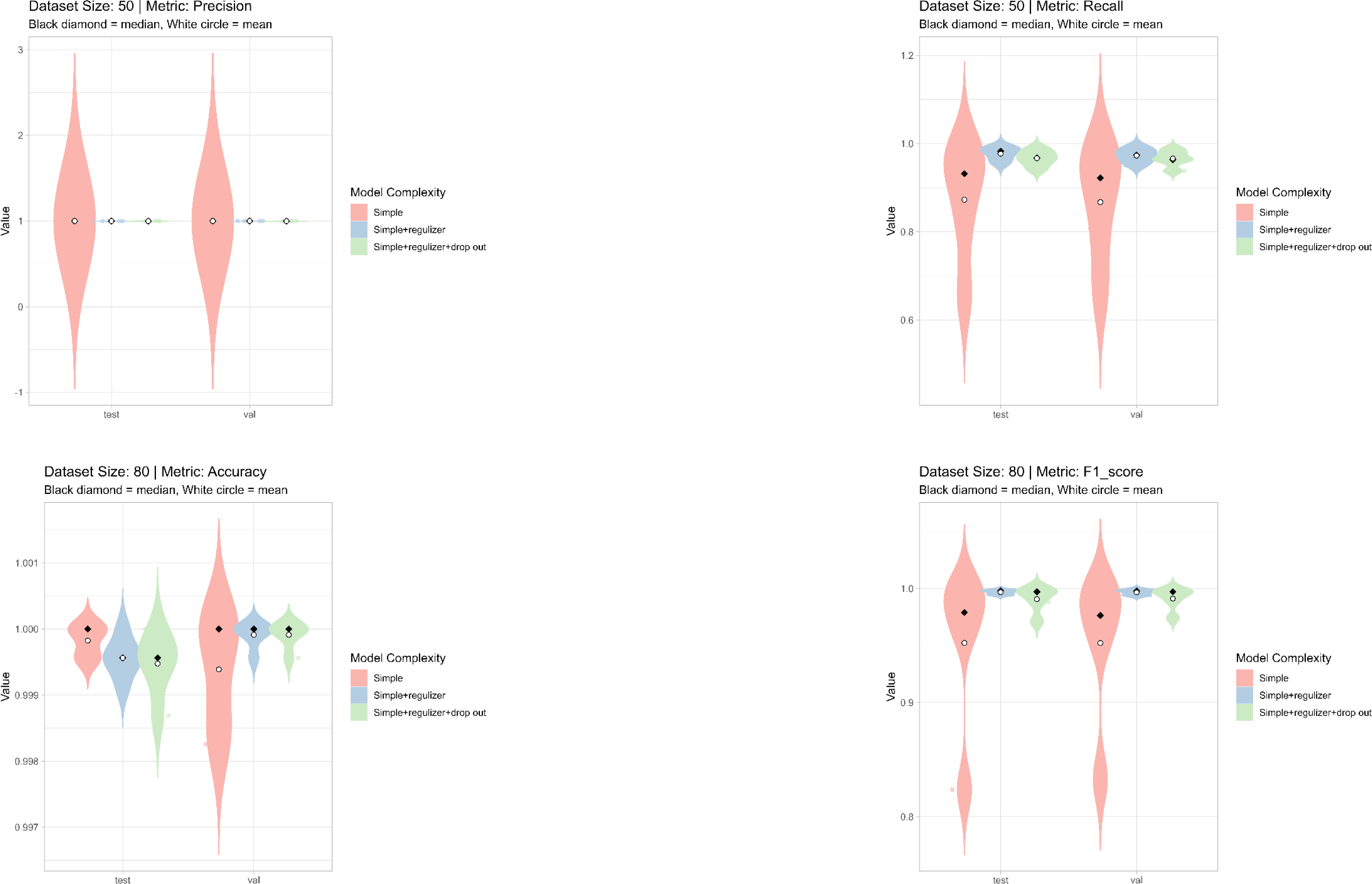

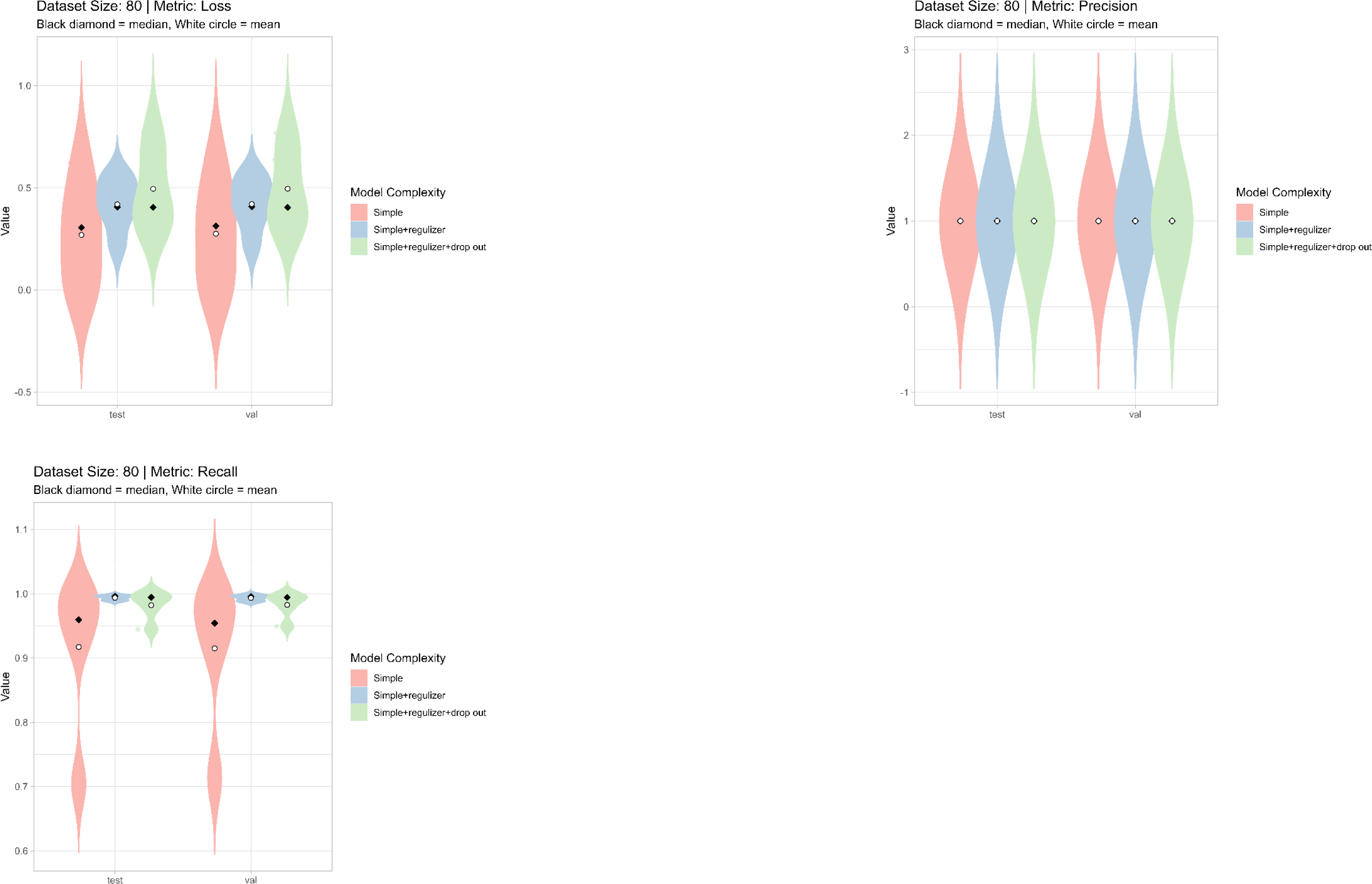

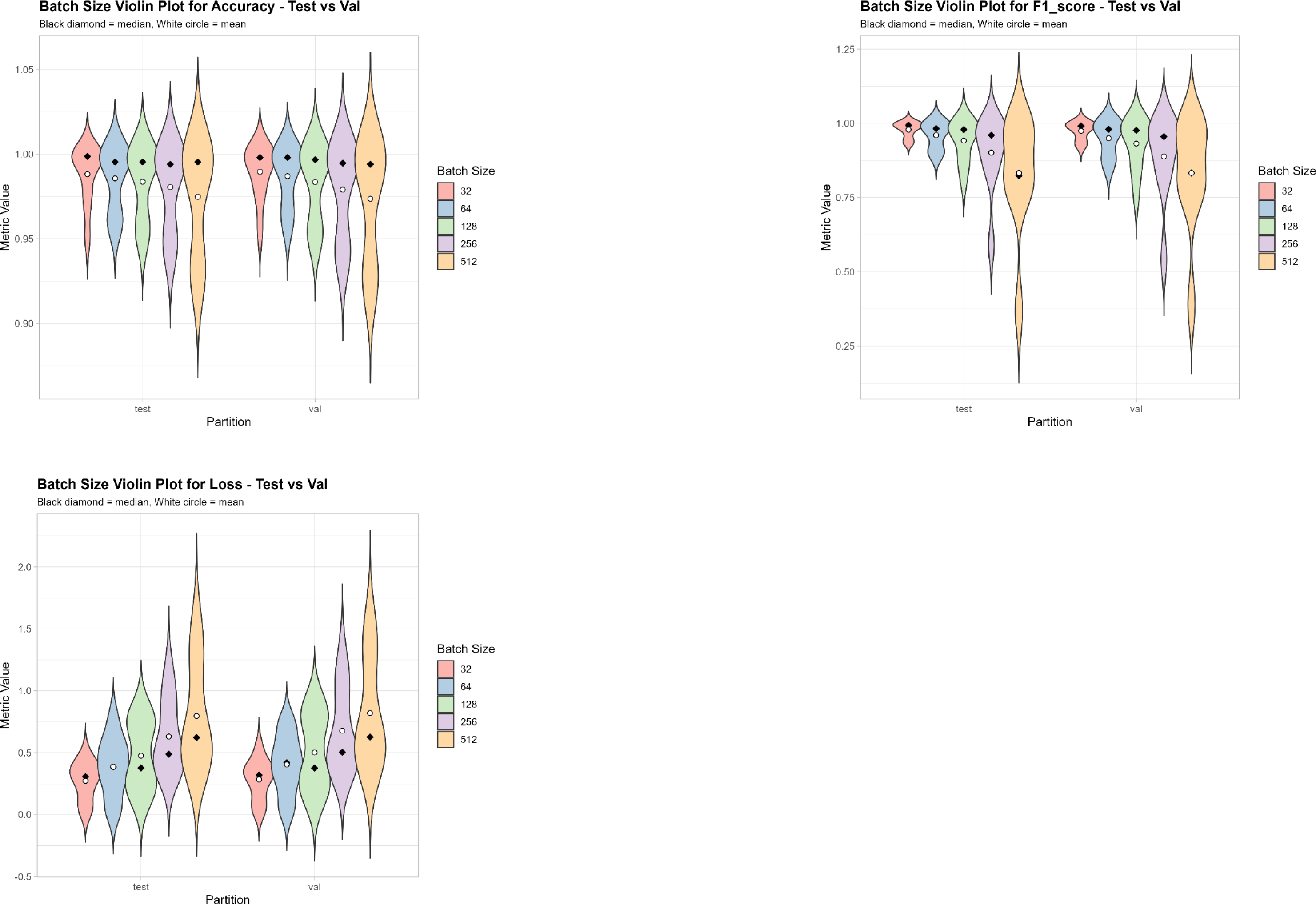

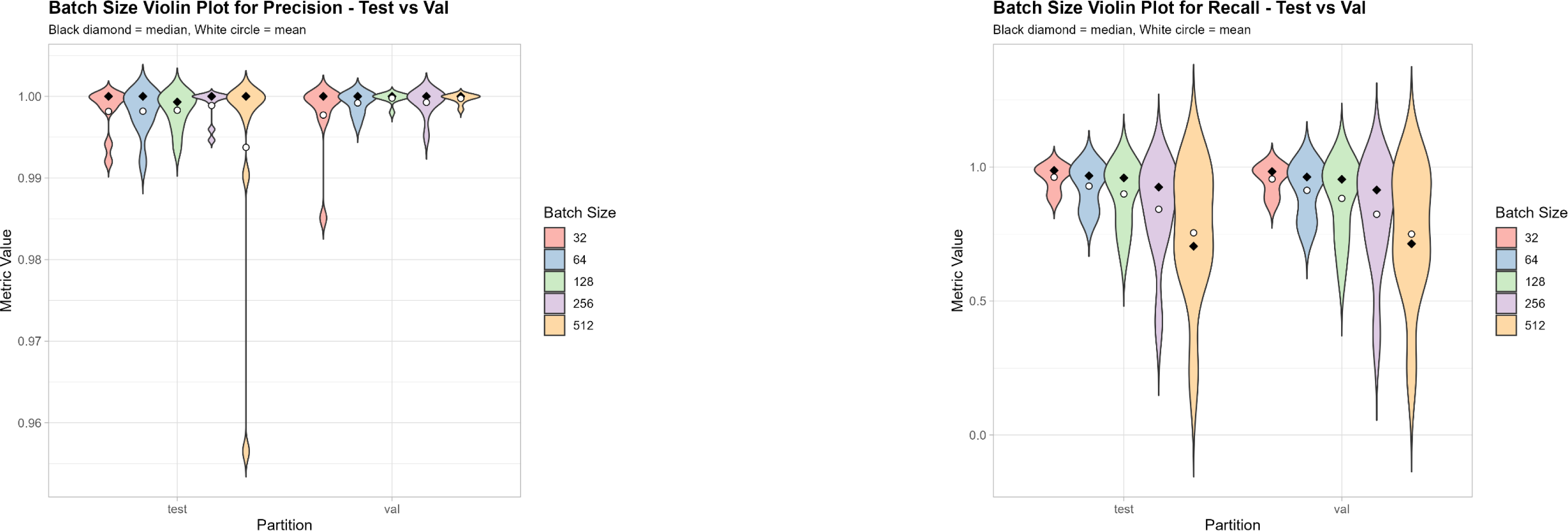

**Data S1 Ablation study with ConvNeXtTiny backbone**

Violin plots visualizing test and validation accuracy, f1_score, recall, precision and loss

Iteration over 20, 50 and 80 augmented images, and simulated over three batch sizes (32, 64, 128)

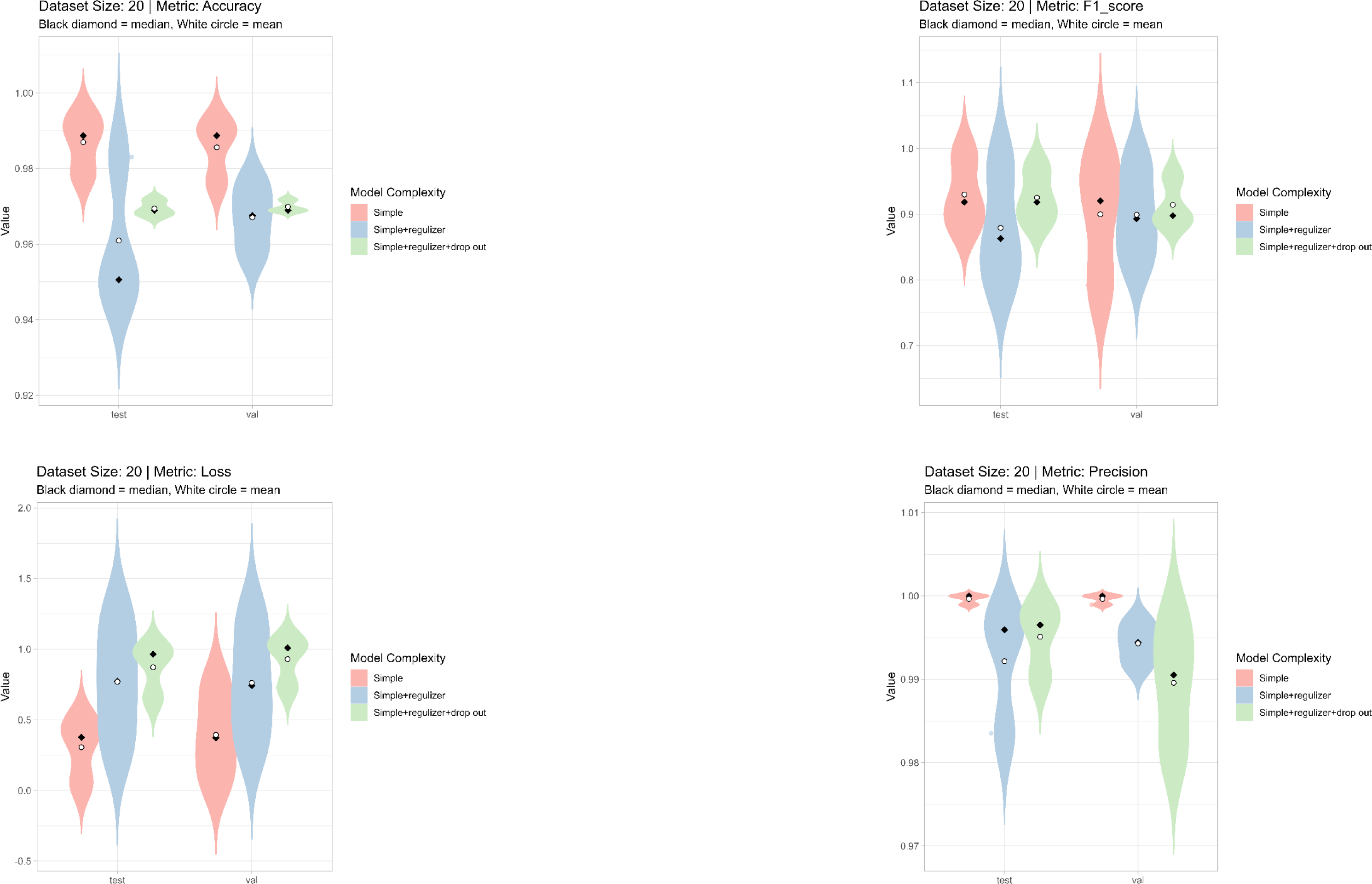

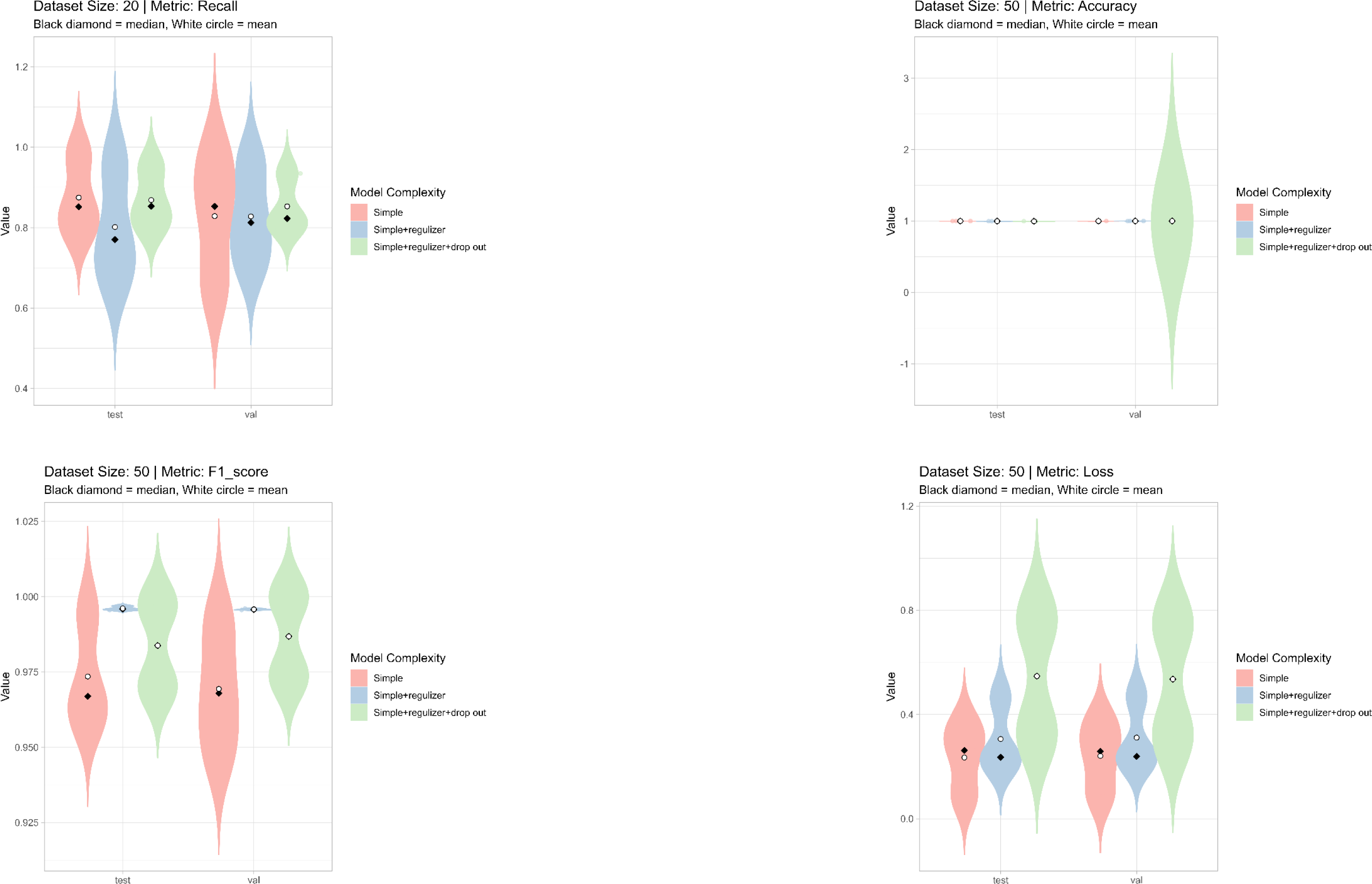

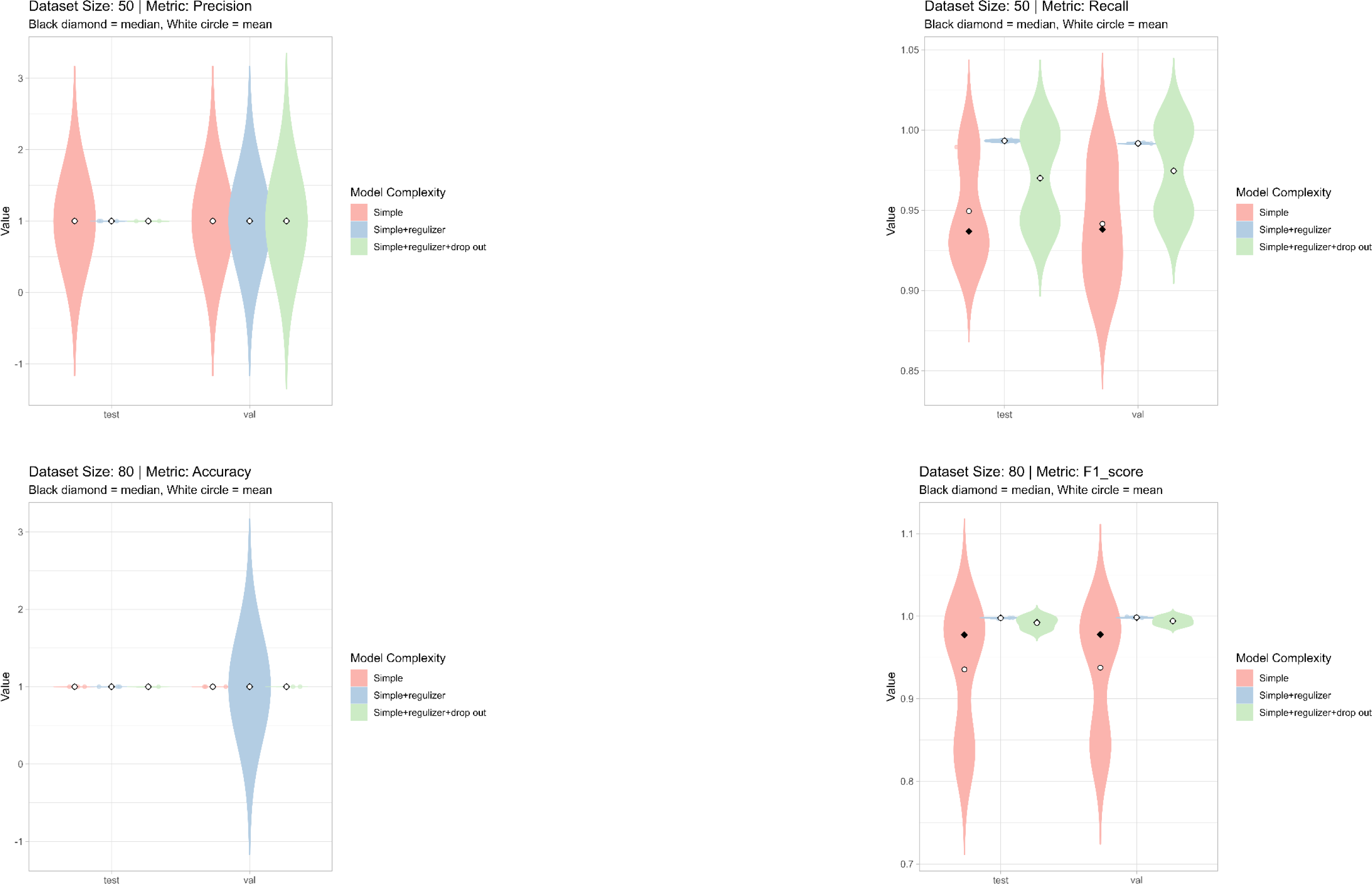

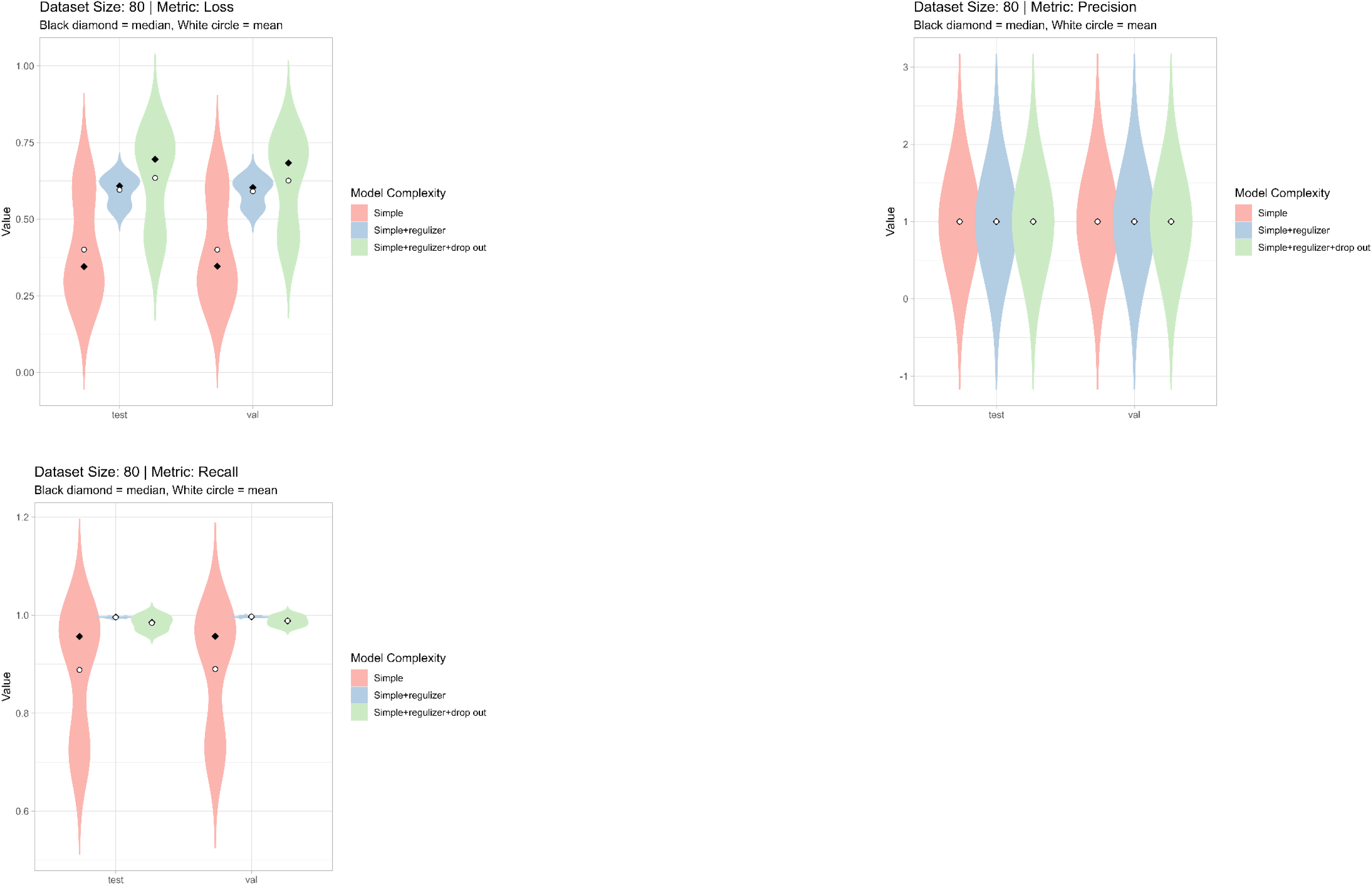

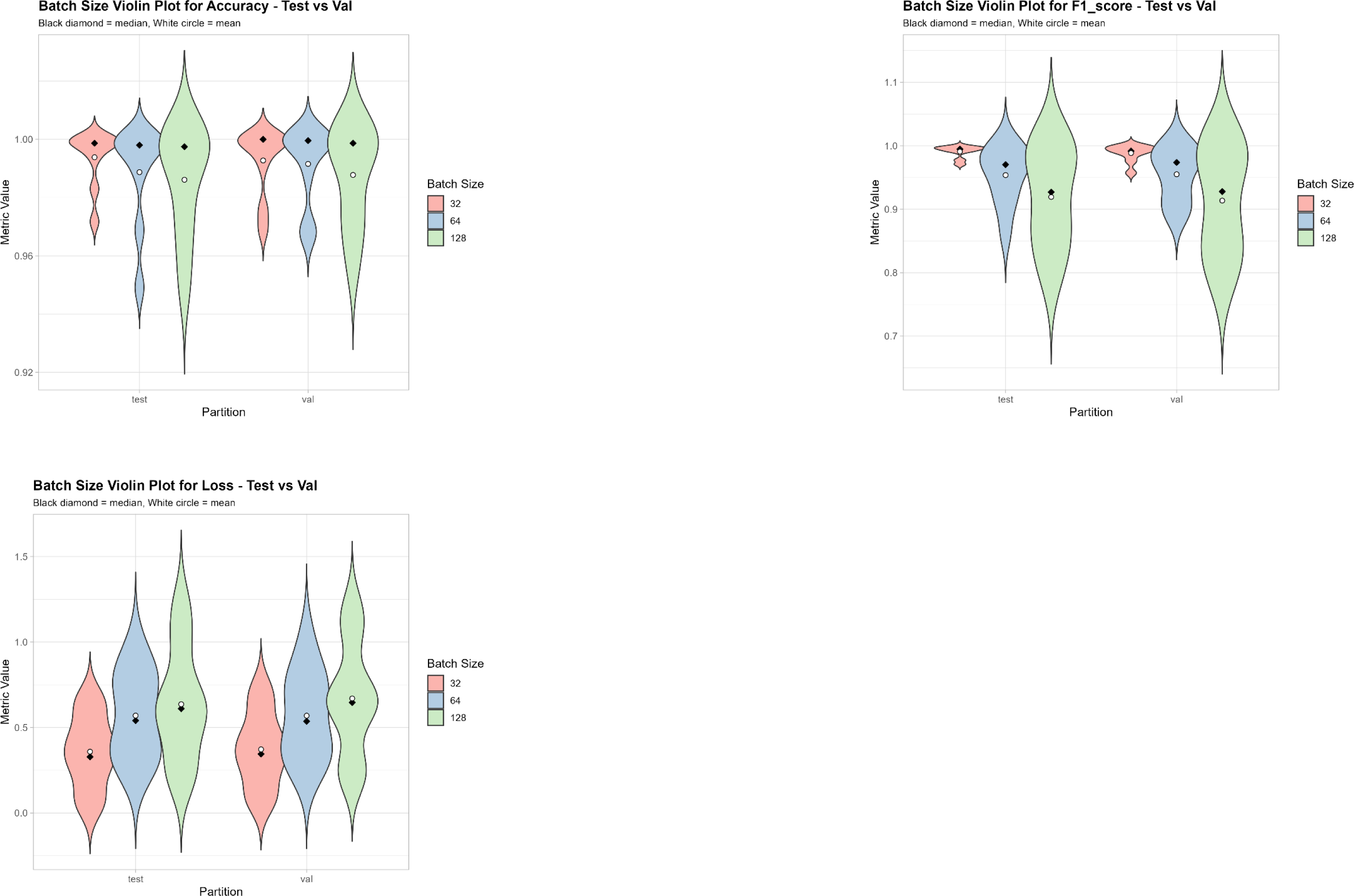

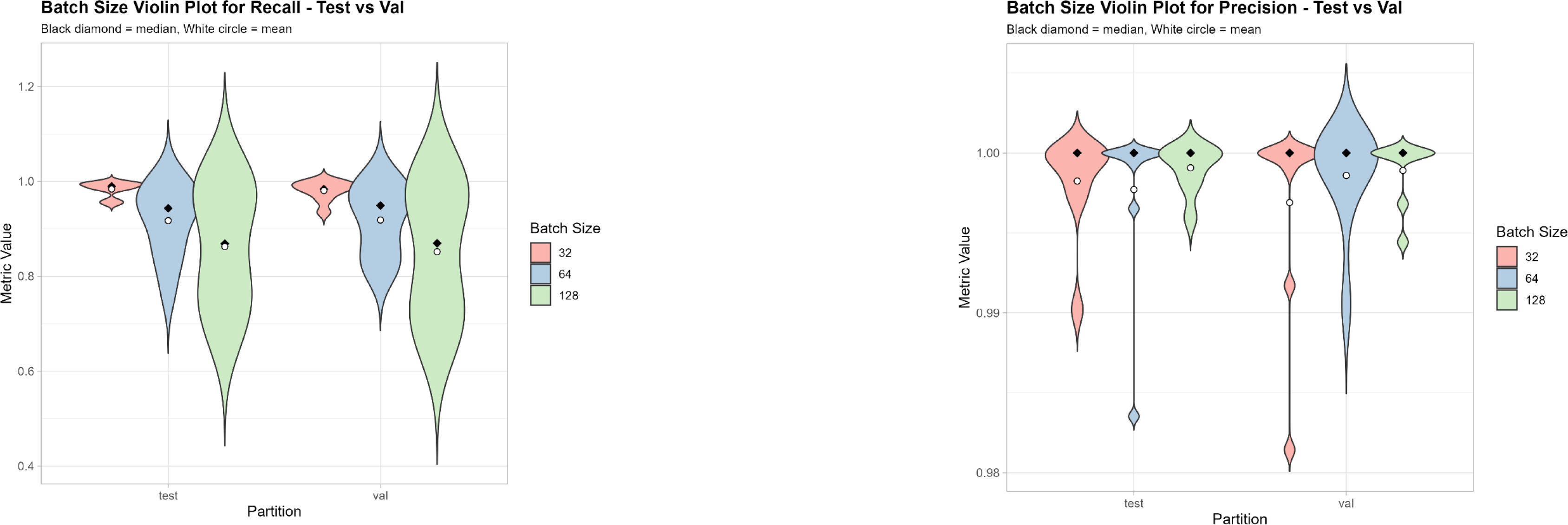

**Data S1 Control study with ConvNeXtTiny backbone on NeuroSYS AGAR**

Violin plots visualizing test and validation accuracy, f1_score, recall, precision and loss

Iteration over 20 augmented images, and simulated over five batch sizes (32, 64, 128, 256, 512)

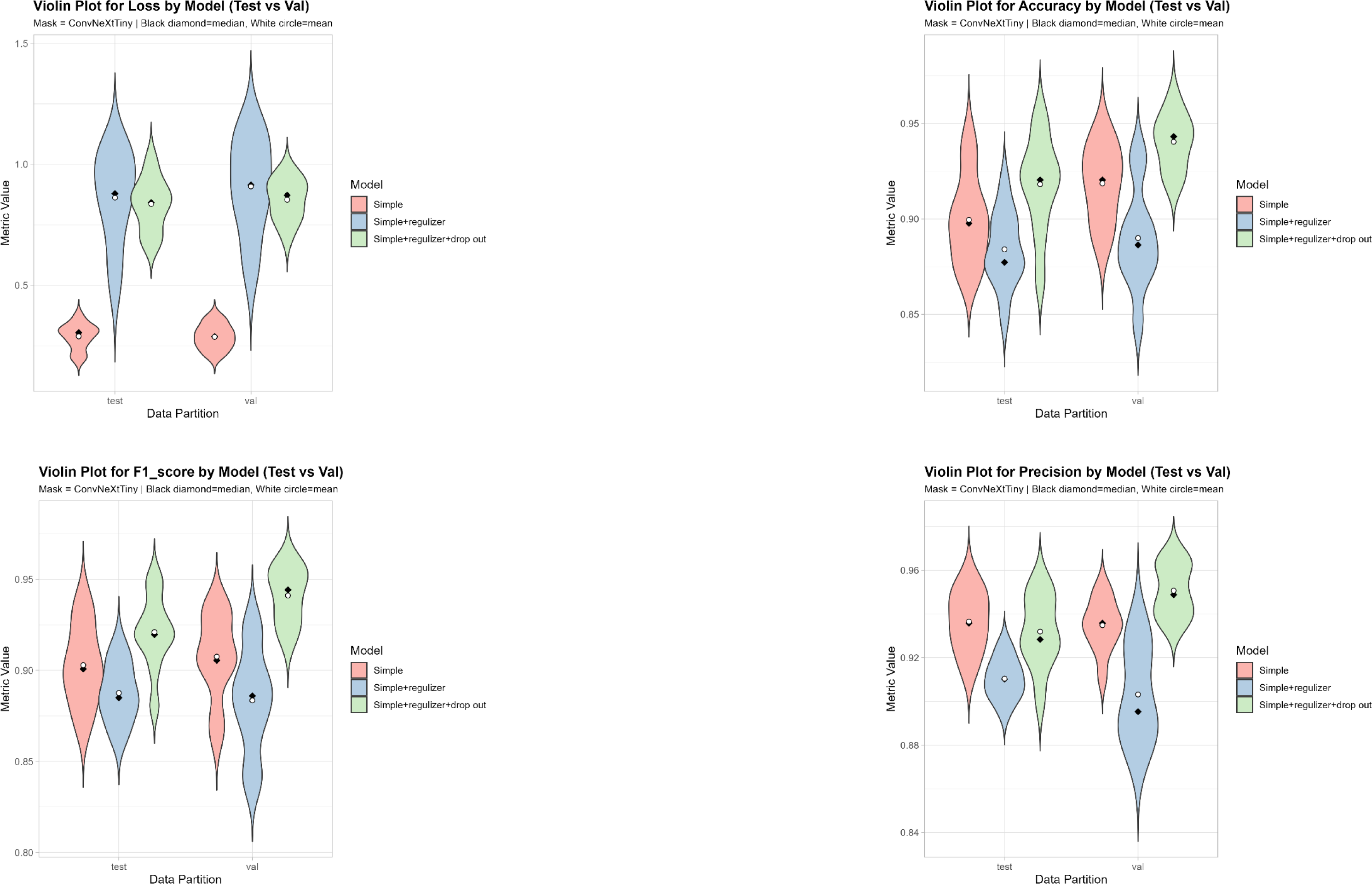

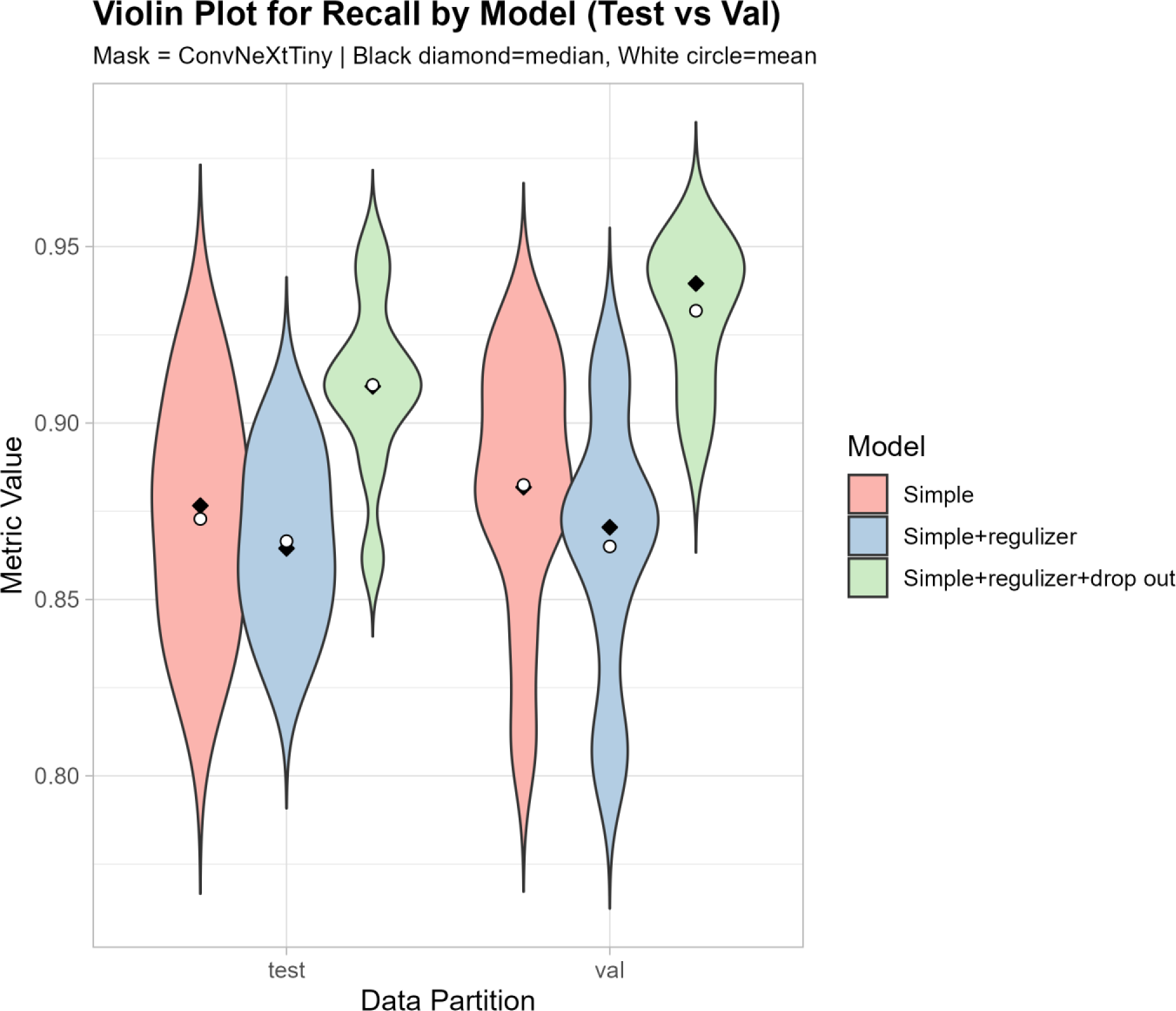

**Data S1 Control study with VGG 16 backbone on NeuroSYS AGAR**

Violin plots visualizing test and validation accuracy, f1_score, recall, precision and loss

Iteration over 20 augmented images, and simulated over five batch sizes (32, 64, 128, 256, 512)

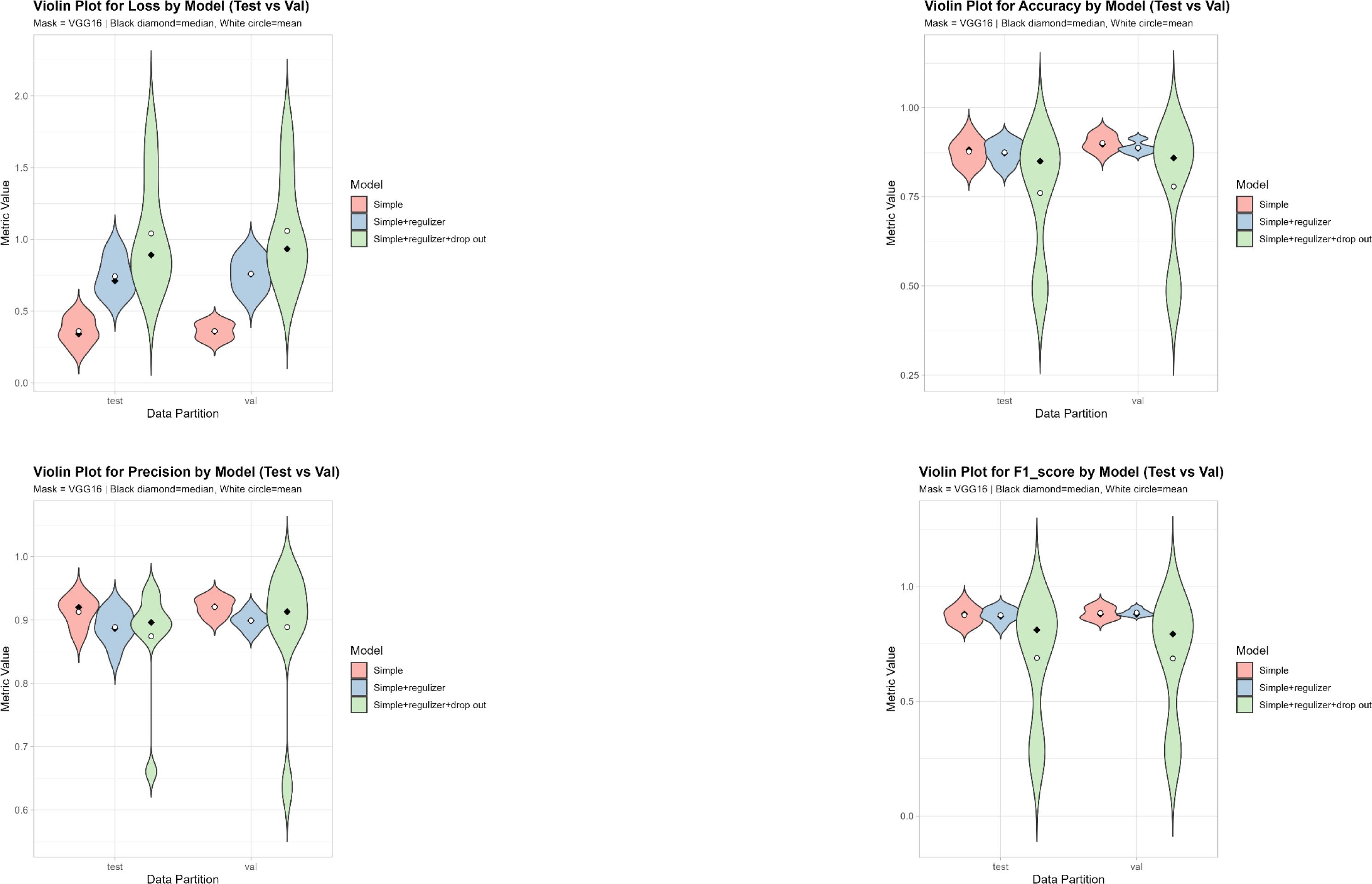

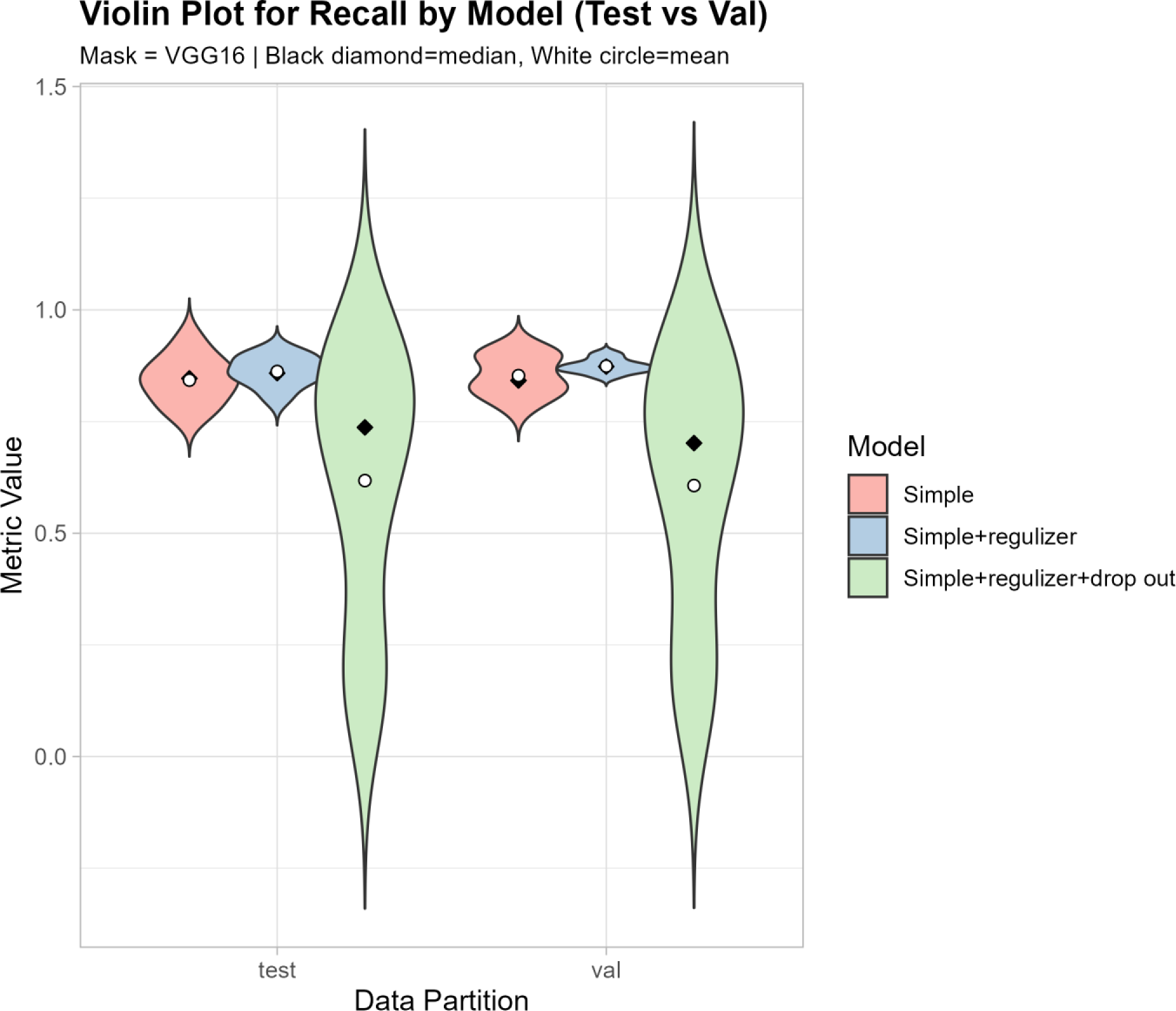

**Data S1 Final Classifier Performance Metrics**

The following slides showcase the performance metrics of the final classifier design, as of figure 2.

The underlying data depicted in figure 2, is the one highlighted for each backbone in the respective table.

**Data S1 Classifier performance with the Densenet_121 backbone**

Metrics over training and validation accuracy, f1_score, recall, precision and loss

Iteration over 40 augmented images, and simulated over five batch sizes (16, 32, 64, 128, 256)

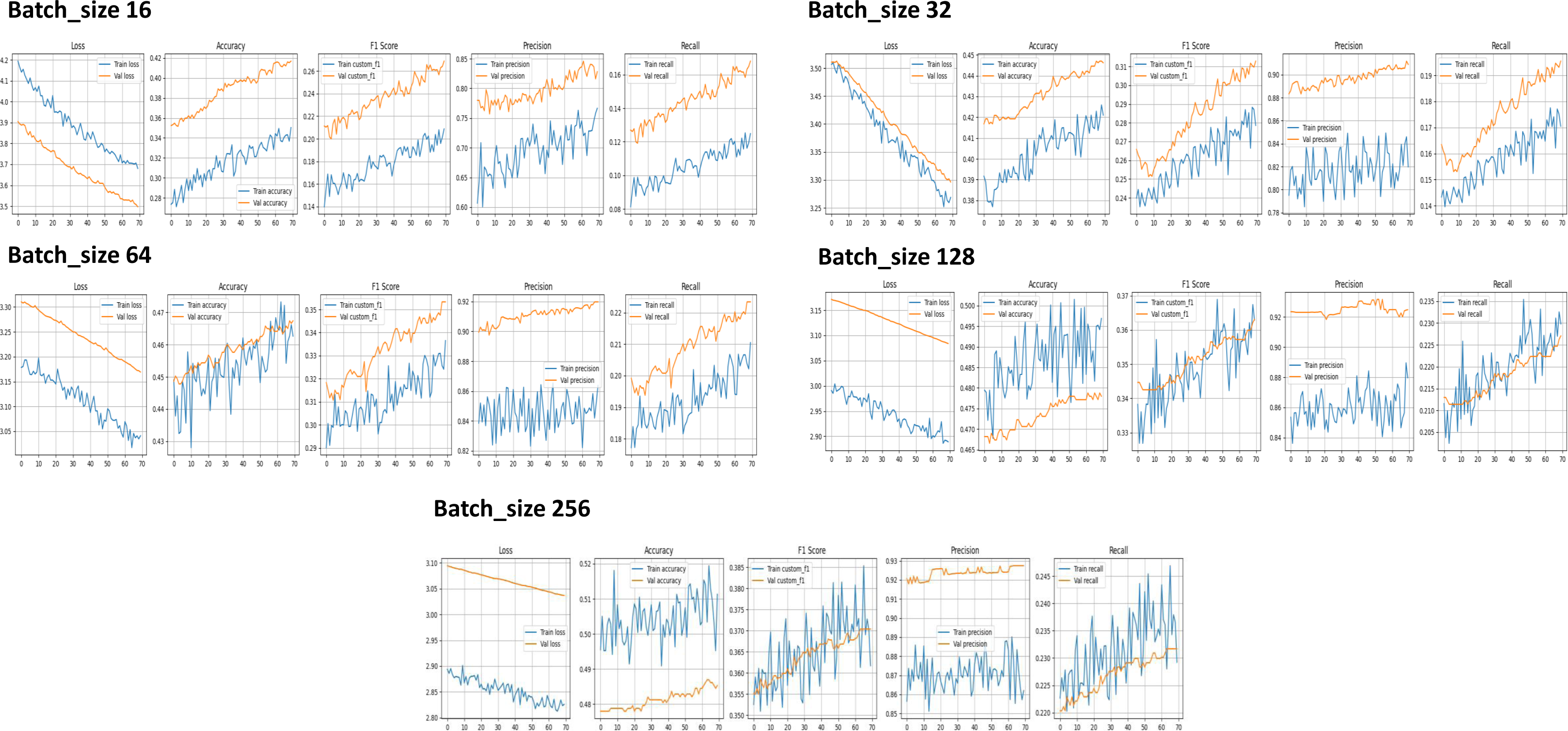

**Data S1 Classifier performance with the Densenet_121 backbone**

Training data

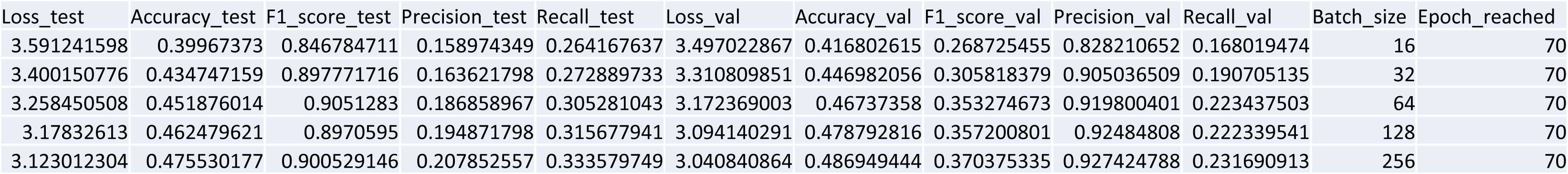

**Data S1 Classifier performance with the Vgg_16 backbone**

Metrics over training and validation accuracy, f1_score, recall, precision and loss

Iteration over 40 augmented images, and simulated over five batch sizes (16, 32, 64, 128, 256)

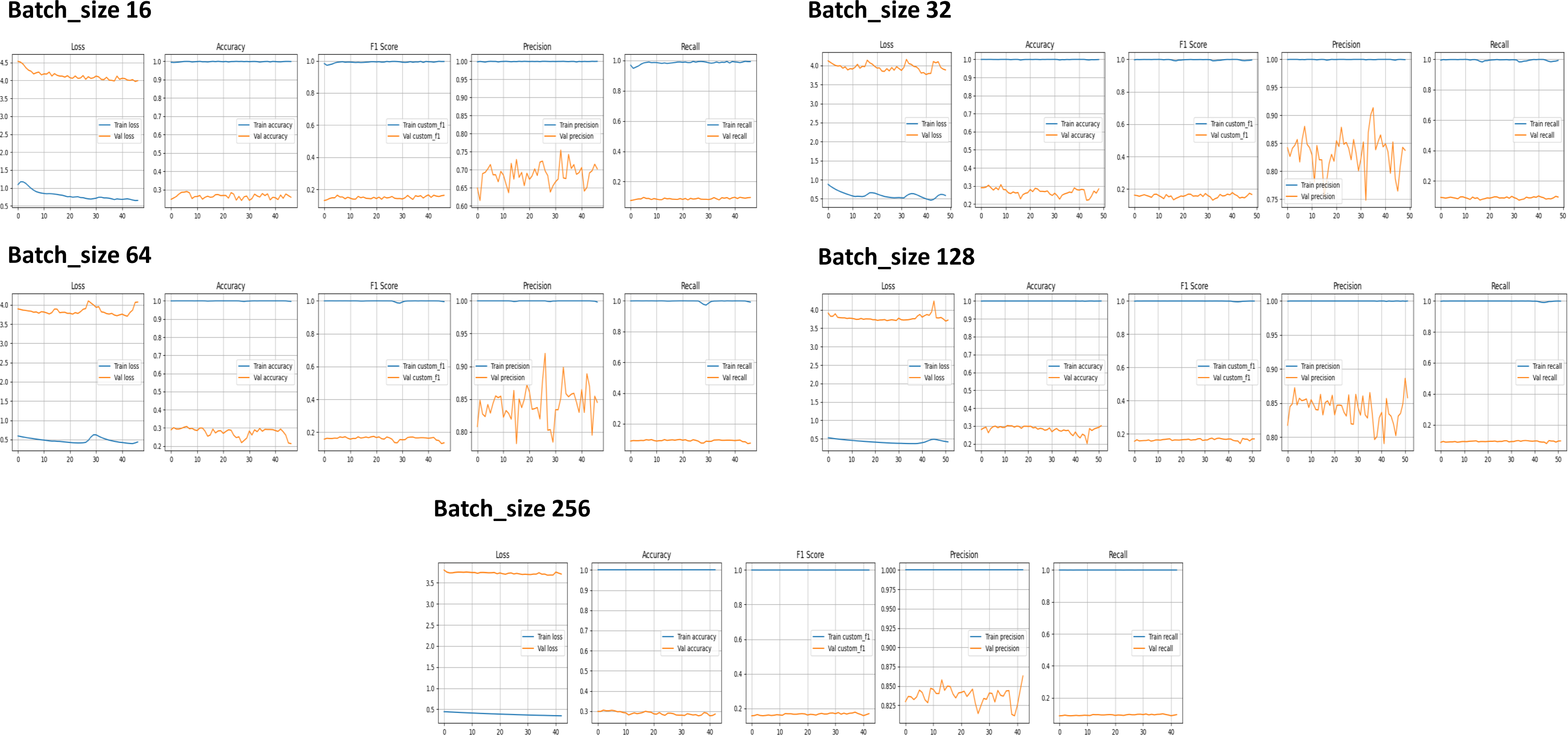

**Data S1 Classifier performance with the Vgg_16 backbone**

Training data

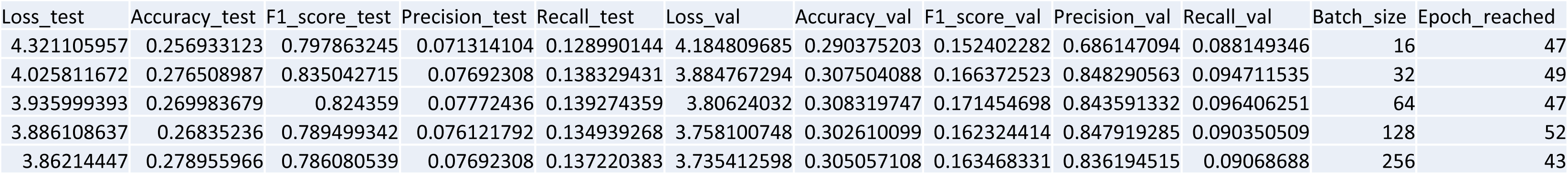

**Data S1 Classifier performance with the Efficientnet_v2b2 backbone**

Metrics over training and validation accuracy, f1_score, recall, precision and loss

Iteration over 40 augmented images, and simulated over five batch sizes (16, 32, 64, 128, 256)

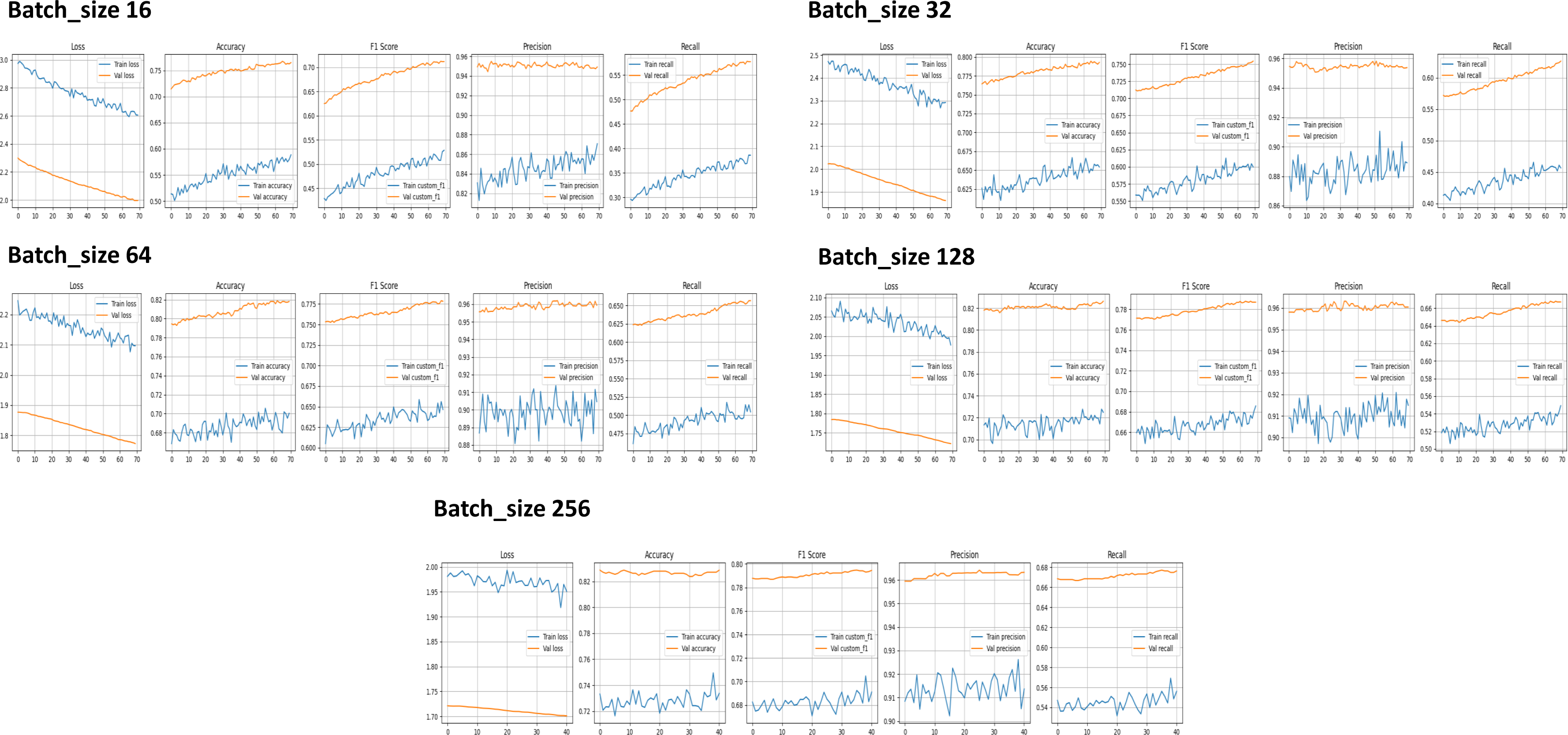

**Data S1 Classifier performance with the Efficientnet_v2b2 backbone**

Training data

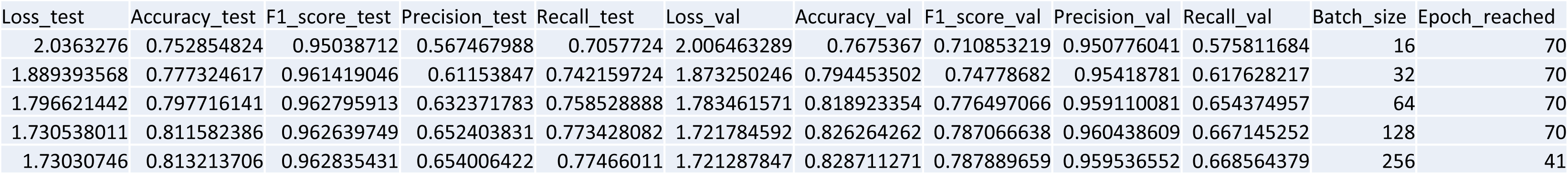

**Data S1 Classifier performance with the ConvNextTiny backbone**

Metrics over training and validation accuracy, f1_score, recall, precision and loss

Iteration over 40 augmented images, and simulated over five batch sizes (16, 32, 64, 128, 256)

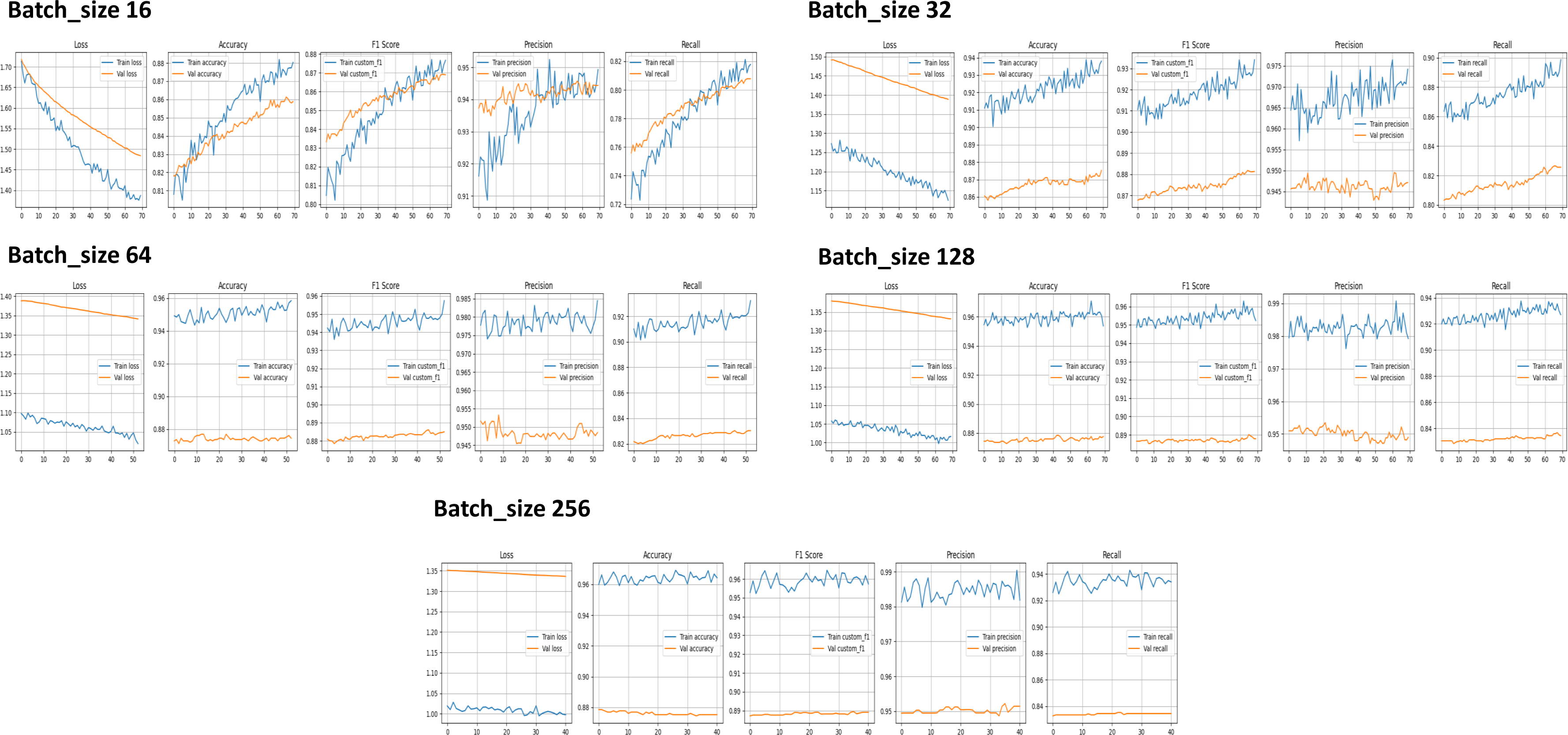

**Data S1 Classifier performance with the ConvNextTiny backbone**

Training data

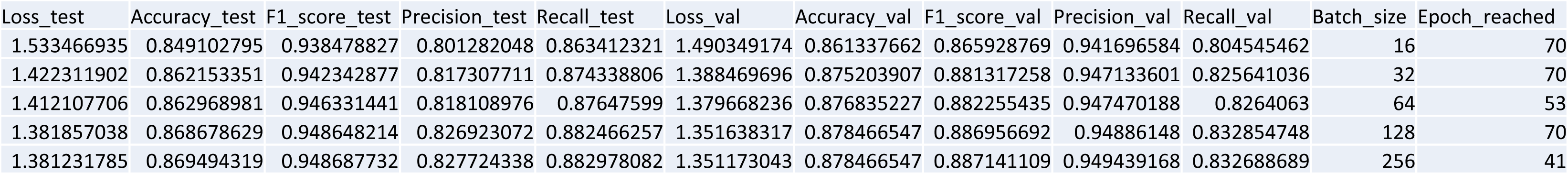

## References

Capper D, Jones DTW, Sill M, Hovestadt V, Schrimpf D, Sturm D, et al. (2018). DNA methylation-based classification of central nervous system tumours. Nature 555:469–474. 10.1038/nature26000

Cheng T, Zhang D, Gu C, Zhou X-G, Qiao H, Guo W, et al. (2024). YOLO-CG-HS: a lightweight spore detection method for wheat airborne fungal pathogens. Comput Electron Agric 227:109544.

Concepcion R II, Guillermo M, Tanner SE, Fonseca V, Duarte B. (2023). BivalveNet: a hybrid deep neural network for common cockle (Cerastoderma edule) geographical traceability based on shell image analysis. Ecol Inform 78:102344. 10.1016/j.ecoinf.2023.102344

Cornely OA, Bassetti M, Calandra T, Garbino J, Kullberg BJ, Lortholary O, et al. (2012). ESCMID guideline for the diagnosis and management of Candida diseases 2012: non-neutropenic adult patients. Clin Microbiol Infect 18 Suppl 7:19–37.

FungiCLEF25 Challenge. (2025). Few-shot learning for fungal identification. Kaggle. https://www.kaggle.com/competitions/fungi-clef-2025/

Giannella M, Lanternier F, Dellière S, Groll AH, Mueller NJ, Alastruey-Izquierdo A, et al. (2025). Invasive fungal disease in the immunocompromised host: changing epidemiology, new antifungal therapies, and management challenges. Clin Microbiol Infect 31:29–36.

Graczyk KM, Pawłowski J, Majchrowska S, Golan T. (2022). Self-normalized density map (SNDM) for counting microbiological objects. Sci Rep 12:10583. 10.1038/s41598-022-14879-3

Gumus A. (2024). Classification of microscopic fungi images using vision transformers for enhanced detection of fungal infections. Turk J Nat Sci 13(1):152–160. 10.46810/tdfd.1442556

Halder A, Gharami S, Sadhu P, Singh PK, Woźniak M, Ijaz MF. (2024). Implementing vision transformer for classifying 2D biomedical images. Sci Rep 14:12567. 10.1038/s41598-024-63094-9

Hörst F, Rempe M, Heine L, Seibold C, Keyl J, Baldini G, et al. (2024). CellViT: Vision transformers for precise cell segmentation and classification. Med Image Anal 94:103143.

Huang T-S, Wang K, Ye X-Y, Chen C-S, Chang F-C. (2023). Attention-guided transfer learning for identification of filamentous fungi encountered in the clinical laboratory. Microbiol Spectr 11(3):e04611–22. 10.1128/spectrum.04611-22

Kiel S. (2021). Assessing bivalve phylogeny using deep learning and computer vision approaches. bioRxiv 2021.04.08.438943. 10.1101/2021.04.08.438943

Krivek G, Gillert A, Harder M, Fritze M, Frankowski K, Timm L, et al. (2023). BatNet: a deep learning-based tool for automated bat species identification from camera trap images. Remote Sens Ecol Conserv 9(6):759–74. 10.1002/rse2.339

Liu Z, Mao H, Wu C-Y, Feichtenhofer C, Darrell T, Xie S. (2022). ConvNeXt: Revisiting ConvNets for 21st century computer vision. arXiv 2201.03545. https://arxiv.org/abs/2201.03545

Ma H, Yang J, Chen X, Jiang X, Su Y, Qiao S, et al. (2021). Deep convolutional neural network: a novel approach for the detection of Aspergillus fungi via stereomicroscopy. J Microbiol 59(6):563–572. 10.1007/s12275-021-1013-z

Microsoft. (2025). The path to Medical Superintelligence. Microsoft AI. https://microsoft.ai/new/the-path-to-medical-superintelligence/

Miao Z, Gaynor KM, Wang J, Liu Z, Muellerklein O, Norouzzadeh MS, et al. (2019). Insights and approaches using deep learning to classify wildlife. Sci Rep 9(1):1–9. 10.1038/s41598-019-51599-8

Pawłowski J, Majchrowska S, Golan T. (2022). Generation of microbial colonies dataset with deep learning style transfer. Sci Rep 12:5212. 10.1038/s41598-022-09264-z

Raghavan K, Sivaselvan B, Kamakoti V. (2024). Attention guided Grad-CAM: an improved explainable artificial intelligence model for infrared breast cancer detection. Multimed Tools Appl 83:57551–78. 10.1007/s11042-023-15500-6

Rani P, et al. (2025). Time-lapse imaging and AI for rapid fungal species identification. arXiv 2501.02855. https://arxiv.org/abs/2501.02855

Rawat S, Bisht B, Bisht V, Rawat N, Rawat A. (2024). MeFunX: a novel meta-learning-based deep learning architecture to detect fungal infection directly from microscopic images. Franklin Open 6:100069.

Selvaraju RR, Cogswell M, Das A, Vedantam R, Parikh D, Batra D. (2020). Grad-CAM: visual explanations from deep networks via gradient-based localization. Int J Comput Vis 128:336–359. 10.1007/s11263-019-01228-7

Shorten C, Khoshgoftaar TM. (2019). A survey on image data augmentation for deep learning. J Big Data 6:60. 10.1186/s40537-019-0197-0

Stiller S, Dueñas JF, Hempel S, Rillig MC, Ryo M. (2024). Deep learning image analysis for filamentous fungi taxonomic classification: dealing with small datasets with class imbalance and hierarchical grouping. Biol Methods Protoc bpae063. 10.1093/biomethods/bpae063

Thirunavukarasu R, Kotei E. (2024). A comprehensive review on transformer networks for natural and medical image analysis. Comput Sci Rev 53:100648.

Torres-Sangiao E, Leal Rodriguez C, García-Riestra C. (2021). Application and perspectives of MALDI–TOF mass spectrometry in clinical microbiology laboratories. Microorganisms 9(7):1539. 10.3390/microorganisms9071539

Tsang CC, Zhao C, Liu Y, Lin KPK, Tang JYM, Cheng KO, et al. (2025). Automatic identification of clinically important Aspergillus species by artificial intelligence-based image recognition: proof-of-concept study. Emerg Microbes Infect 14(1):2434573. 10.1080/22221751.2024.2434573

Weber S, et al. (2023). FUTURE-AI: A framework for trustworthy AI in healthcare. arXiv 2309.12325. https://arxiv.org/abs/2309.12325

Wu Y, Gadsden SA. (2023). Machine learning algorithms in microbial classification: a comparative analysis. Front Artif Intell 6:1200994. 10.3389/frai.2023.1200994

Yang F, Zhong Y, Yang H, Wan Y, Hu Z, Peng S. (2023). Microbial colony detection based on deep learning. Appl Sci 13(19):10568. 10.3390/app131910568

Yang S, Xiao W, Zhang M, Guo S, Zhao J, Shen F. (2023). Image data augmentation for deep learning: a survey. arXiv 2204.08610. https://arxiv.org/abs/2204.08610

Zhang L, et al. (in press 2025). Fusion of MALDI-ToF spectra with colony imagery via dual-encoder networks. J Clin Mycol.

